# First quarter chronicle of COVID-19: an attempt to measure governments’ response

**DOI:** 10.1101/2020.09.20.20198242

**Authors:** Şule Şahin, María del Carmen Boado-Penas, Corina Constantinescu, Julia Eisenberg, Kira Henshaw, Maoqi Hu, Jing Wang, Wei Zhu

## Abstract

The crisis caused by COVID-19 revealed the global unpreparedness for handling the impact of a pandemic. In this paper, we present a first quarter chronicle of COVID-19 in Hubei China, Italy and Spain, specifically their infection speed, death and fatality rates. By fitting distributions to these rates, we look for the effectiveness of government measures during the pandemic through a number of statistical approaches.

## 1 Introduction

First reports of an outbreak of the (2019-nCov)-novel coronavirus-infected pneumonia (NCIP) were identified in December 2019 in the city of Wuhan, Hubei Province, China. Initially identified as pneumonia of unknown origin, confirmed cases were found to bear similarities to the severe acute respiratory syndrome coronavirus (SARS-CoV) of 2003 and the Middle East respiratory syndrome (MERS-CoV) of 2012, with the earliest instances of the virus linked to the Huanan Seafood Wholesale Market in Wuhan. Declared a pandemic by the World Health Organisation on 11 March 2020, the highly contagious virus rapidly became a global concern, now present in more than 200 territories.

According to the World Health Organization (WHO), coronaviruses (CoV) are a large family of viruses that cause illnesses ranging from the common cold to more severe diseases such as SARS-CoV and MERS-CoV. The outbreak of SARS-CoV, Deng and Peng [3], began in China in 2002 and was overcome by disease prevention and control systems. MERS-CoV was first reported in Saudi Arabia in 2012 and has since spread to several other countries. Although most coronavirus infections, Huang et al. [5], Sohrabi et al. [10] and Zhu et al. [17], are not severe, more than 10,000 cumulative cases have been associated with SARS-CoV and MERS-CoV in the past two decades, with mortality rates of 10% and 37% respectively.

For governments, there are two possible strategies for handling an epidemic: a) mitigation, focusing on slowing the epidemic without interrupting transmission completely, i.e. reducing the peak healthcare demand while protecting vulnerable groups, and b) suppression, which aims to reverse epidemic growth through the reduction of the number of cases to a minimal level. Mitigation involves isolating suspected cases, quarantining the households of suspected cases and socially distancing the most vulnerable during the peak of the outbreak. While suppression measures include the social distancing of the entire population with the added possibility of school and university closures. Suppression, although successful to date in China^1^ and South Korea, carries with it enormous social and economic costs which may significantly impact the well-being of society in the short and long run. Hence, there is a trade-off between minimising deaths from a pandemic and the economic impact of viral spread.

In this paper, we aim to provide a thorough discussion of the changing nature of the COVID-19 pandemic during the first three to four months of outbreaks in Hubei, Italy and Spain. All three regions adopted suppression strategies for mitigating the risk of the pandemic, with severe lockdown restrictions governing the corresponding populations. Considering infection speed, death rate and fatality ratio, we observe potential implications of the implementation of government measures. We adopt a data driven approach for the analysis, exploring probability distributions of daily cross-sectional data in each of the three cases, in addition to presenting descriptive statistics for the severity measures.

Distribution fitting provides further indication of how the significance of the outbreak changes throughout the observation period. In order to fit daily distributions, we assume the daily rates for each city in a region or country to be independent of the rates in all other cities and fit distributions to each day. Referring to Table 1, we assume the independence of columns and fit distributions to every row. We then analyse the daily trend in parameters of the most common best fitting distribution, which is fitted to all dates. The process is repeated for all three territories considered, using the distribution fitting software Easyfit for the analysis.

**Table 1:**
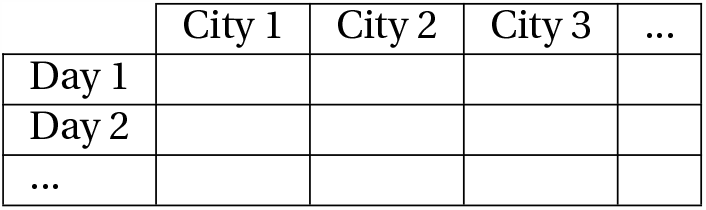
Explanation of the distribution fitting methodology

**Table 2.** Timeline of the COVID-19 pandemic in Hubei (December 2019 - April 2020)

|  |  |
| --- | --- |
| 1 Dec (Day 1) | First case identified |
| 31 Dec (Day 31) | Unknown pneumonia outbreak announced |
| 1 Jan (Day 32) | Huanan Seafood Market closed |
| 9 Jan (Day 40) | First death occurred |
| 20 Jan (Day 51) | Human-to-human transmission confirmed |
| 23 Jan (Day 54) | Lockdown in Wuhan began |
| 28 Jan (Day 59) | Quarantine restrictions governed Hubei province entirely |
| 3 Feb (Day 65) | First speciality field hospital began accepting patients |
| 8 Feb (Day 70) | Second field hospital opened |
| 13 Feb (Day 75) | Hubei reported approximately 10 times more new cases than the previous day due to the new definition of confirmed cases |
| 13 Mar (Day 104) | Easing of restrictions began in Hubei, with the exception of Wuhan |
| 25 Mar (Day 116) | All lockdowns outside Wuhan lifted |
| 9 Apr (Day 131) | Lockdown in Wuhan lifted and all transportation resumed |
| 26 Apr (Day 148) | All coronavirus patients in Wuhan discharged |

**Table 3.** Timeline of the COVID-19 pandemic in Italy (January 2020 - June 2020)

|  |  |
| --- | --- |
| 31 Jan (Day 1) | First two cases confirmed; state of emergency declared |
| 22 Feb (Day 23) | First two deaths occurred; partial lockdowns imposed in two regions in Northern Italy |
| 25 Feb (Day 26) | Cases emerged in other regions |
| 4 Mar (Day 34) | Schools and universities closed nationwide; 100 deaths reached |
| 7 Mar (Day 37) | Lockdown extended to cover more areas |
| 10 Mar (Day 40) | Nationwide lockdown began |
| 21 Mar (Day 51) | All non-essential production, industry and business halted |
| 1 Apr (Day 62) | Lockdown extended until 13 April |
| 8 Apr (Day 69) | All Italian ports closed until 31 July |
| 10 Apr (Day 71) | Lockdown prolonged until 3 May; some businesses reopened |
| 20 Apr (Day 81) | First fall in the number of active cases observed |
| 4 May (Day 95) | Manufacturing industries and construction sites reopened |
| 18 May (Day 109) | Most businesses allowed to reopen; movement granted with each region |
| 25 May (Day 116) | Swimming pools and gyms reopened |
| 15 Jun (Day 137) | Theatres and cinemas reopened |

**Table 4.**
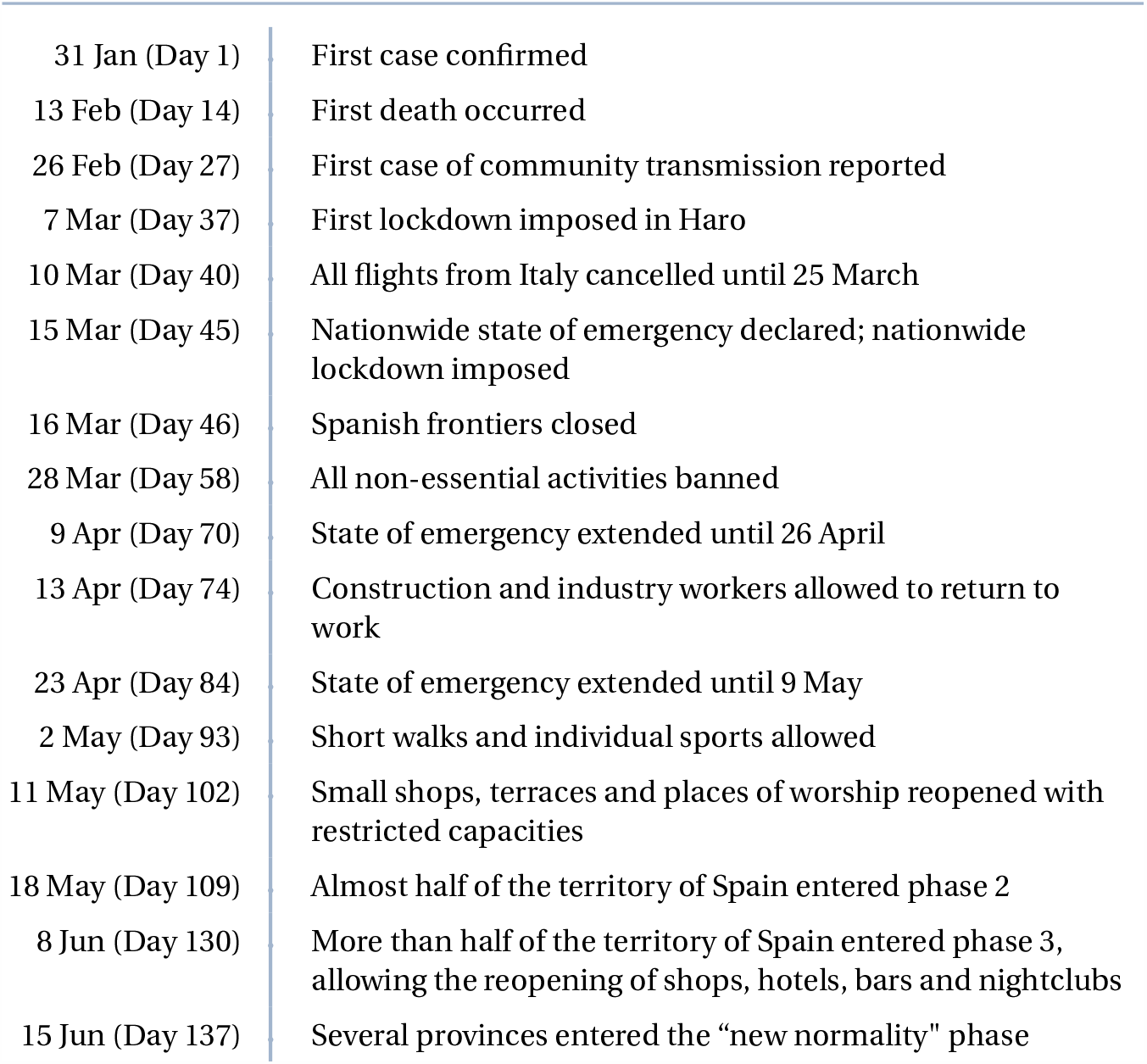
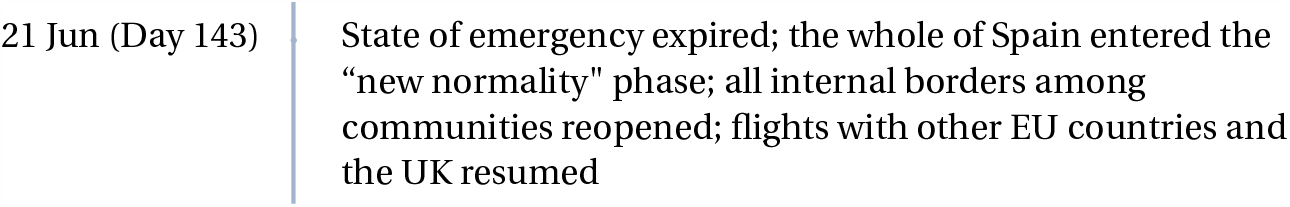
Timeline of the COVID-19 pandemic in Spain (January 2020 - June 2020)

During the initial stages of the pandemic, government measures were generally standardised throughout each region under consideration. We therefore consider the outbreaks in each of Hubei, Italy and Spain as single outbreaks, rather than analysing the data at the more granular city level. However, as local lockdowns become more common, with increases in the number of cases occurring in particular regions, analysis of the nature of the pandemic in the period after our observation may benefit from the breaking down of regions into cities, so as not to temper the significance of any local outbreak.

A number of limitations in regard to the data sets used for the observations in this paper include issues with the timely reporting of deaths, testing capacities and rates per region, reporting reliability and the non-uniform definition of COVID-19 deaths. Our analysis however provides sufficient insight into the change in the severity of the COVID-19 pandemic over time and the potential implications of lockdown measures.

The remainder of the paper is structured as follows. In Section 2 we present the methodology for the analysis, with particular focus on definition of the three severity measures. Sections 3, 4 and 5 present the COVID-19 timeline, descriptive analysis and distribution fitting for each of Hubei, Italy and Spain, respectively. We provide the concluding remarks of the paper in Section 6.

## 2 Methodology

### 2.1 Three measures of severity

Throughout the paper, we use the notation CC_*t*_, CD_*t*_ and CR_*t*_ to denote the number of cumulative confirmed cases, cumulative deaths and cumulative recovered cases respectively at time *t*. Our analysis focuses on three variables, infection speed, death rate and fatality ratio. The infection speed for a city at time *t, v*_*t*_, is defined by the division of newly confirmed cases at time *t* (cumulative confirmed cases at time *t* − 1 subtracted from cumulative confirmed cases at time *t*) by currently uninfected cases at time *t*−1 (cumulative confirmed cases at time *t*−1 subtracted from total population (TP)), such that

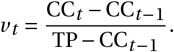

The death rate for a city at time *t*, Δ_*t*_, is defined by the division of new deaths at time *t* (cumulative deaths at time *t* − 1 subtracted from cumulative deaths at time *t*) by currently infected cases at time *t* − 1 (sum of cumulative deaths and recovered cases at time *t* − 1 subtracted from cumulative confirmed cases at time *t* − 1), such that

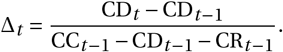

Finally, the fatality rate for a city at time *t, ψ*_*t*_, is defined by the division of cumulative deaths by cumulative confirmed cases at time *t*, such that

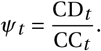

We make two important assumptions. Firstly, daily updated population data is unavailable, we therefore obtain Hubei population data from the Hubei Statistical Yearbook 2019, published by the Hubei Provincial Bureau of Statistics [6]. Italian population data is taken from the Italian National Institute of Statistics [8] and Spanish population data is from the Spain National Statistics Institute [11]. Secondly, we assume individuals who have recovered from the virus to be immune to future infection, and so do not include recovered cases in calculation of the infection speed. The second assumption does not, in fact, significantly affect the analysis, since the number of recovered cases is negligible in comparison to the total population.

### 2.2 Distribution fitting

Following definition of the infection speed, death rate and fatality ratio, daily rates for each city in Hubei and each region in Italy and Spain were calculated. A total of 61 distributions were fitted to these rates on every day of the observation period. Based on Kolmogorov-Smirnov statistics, the best three performing distributions were identified for each day. The distribution most frequently outperforming all alternative distributions was selected to be fitted to all dates in the period. In this analysis, we aim to provide an overview of trends in the nature of the pandemic in Hubei, Italy and Spain, observing potential indications of the effectiveness of government measures.

Distribution fitting results for each of the three territories are presented alongside descriptive statistics of the severity measures in Sections 3, 4 and 5, respectively.

## 3 Hubei, China

### 3.1 Timeline of events

We now present a timeline of COVID-19 events in Hubei, from 1 December 2019 to 26 April 2020.

#### Prior to the “whistleblowing” (1 December - 30 December)

With “patient zero” yet to be confirmed ^2^, it was commonly agreed that the first COVID-19 patient was identified at Jin Yin-tan Hospital, Wuhan, China, on 1 December 2019, with no epidemiological link to later cases [5]. [9] declare that human-to-human transmission has occurred among close contacts since the middle of December 2019. On 30 December, Wuhan ophthalmologist Dr. Li Wenliang, the whistleblower, sent a message to fellow doctors in an online chat group disclosing the occurrence of 7 cases of SARS at the Wuhan Central Hospital, each connected to the Huanan Seafood Wholesale Market ^3^. The doctor provided a warning to colleagues to wear protective clothing in order to avoid infection ^4^. On the evening of 30 December, the Wuhan Municipal Health Commission sent hard-copy messages to its affiliated institutions, containing guidelines on confronting a possible outbreak of infectious pneumonia ^5^.

#### Community transmission (31 December - 22 January)

On 31 December, the Wuhan Municipal Health Commission released an online briefing regarding the pneumonia outbreak in the city ^6^. Information reached the World Health Organisation (WHO) through official channels the same day ^7^. According to the Chinese state-sponsored Xinhua News, the Huanan Seafood Market was closed on 1 January 2020 for “remediation” ^8^. On 9 January, the first death due to the virus occurred in a 61-year-old man. Dying of heart failure and pneumonia, the individual was a regular customer at the market with several significant medical conditions, including chronic liver disease ^9 10^. On 11 January, the WHO published initial guidance on travel advice, testing in laboratories and medical investigations. On 18 January, the Wuhan City government held an annual banquet in the Baibuting community to celebrate Chinese New Year, with forty thousand families in attendance sharing meals, plates and eating together. On 20 January, following the infection of two medical staff in Guangdong, the China National Health Commission confirmed the virus to be human-to-human transmissible ^11^.

#### Lockdown and quarantine restrictions (23 January - 10 March)

At 2 am on 23 January, authorities issued a notice informing residents of Wuhan that from 10 am, all public transport, including buses, railways, flights, and ferry services would be suspended. Wuhan Airport, Wuhan railway station, and Wuhan Metro were all closed. Residents of Wuhan were not allowed to leave the city without permission from authorities. Travel quarantine in Wuhan delayed the overall epidemic progression by just 3 to 5 days in Mainland China, but had a more marked effect on an international scale, where case importations were reduced by nearly 80% until mid February [1]. A report published in The Lancet on 24 January indicated that individuals may be symptom-free for several days while the coronavirus is incubating, increasing the risk of contagious infection without forewarning signs [5]. On 24 January, travel restrictions enacted in 12 additional prefecture-level cities in Hubei meant that apart from 2 prefecture-level districts, the entire Hubei province was under a city-by-city quarantine. On 28 January Xiangyang was the final city in Hubei to become quarantined. On 2 February, Huoshenshan Hospital, an emergency specialty field hospital in Wuhan, was handed over to the People’s Liberation Army after 10 days’ construction undertaken since the first day of lockdown in the city. The hospital began accepting patients the next day ^12^. Applying the same design, a second field hospital, Leishenshan Hospital, opened on 8 February ^13^. On 9 February, the death toll in China surpassed that of the 2002-2003 SARS epidemic, with 811 deaths recorded. On 13 February, Hubei reported 14,840 newly confirmed cases, nearly 10 times more than the previous day, while deaths more than doubled, reaching 242. This large increase was due to a change in the definition of confirmed cases to include clinical diagnoses of patients ^14^. On the same day, the Chinese government issued an extension of order to shut down all non-essential companies, including manufacturing plants, and all schools in Hubei province until 20 February, which was later further extended to 10 March.

#### Easing of lockdown (11 March - 7 April)

From 13 March, with the exception of Wuhan, cities in Hubei province gradually began to remove controls and permits on road traffic within their urban areas. China reported no new locally transmitted infections for the first time since the pandemic began. On 25 March, Hubei lifted the lockdown outside of Wuhan ^15^ and on 8 April, Wuhan lifted its lockdown and resumed all methods of transportation ^16^.

#### Normality (8 April -)

On 17 April, Wuhan revised its official death toll due to the coronavirus from 1,290 to 3,869; with local authorities citing incorrect reporting, delays, and omissions as reasons for the change ^17 18^. On 26 April, the National Health Commission reported that all coronavirus patients in Wuhan had been discharged from the city’s hospitals

### 3.2 Data

The R package, nCov2019, developed by Yu [16], provides direct access to real-time epidemiological data on the outbreak. There are two kinds of data available from this package, real-time data and historical data. The real-time data, which contains current numbers of confirmed cases and deaths in geographical locations, are retrieved using API (application programming interface) calls to the Tencent SARS-COV-2 website [12]. The Tencent website relies on the data obtained from the Chinese provincial health agencies, the China National Health Commission (CNHC), the World Health Organisation (WHO) and public health agencies in other countries.

The historical data provided by the package, which forms the basis of our analysis for Hubei province, has three different sources. The first source is obtained directly from the CNHC^20^, the second source is a non-governmental organisation Dingxi- angyuan ^21^ and the third is a public GitHub repository. ^22^ All three historical datasets are updated daily and are almost consistent with one other [13]. We choose the third source since it provides the earliest data.

The data offers a unique opportunity to study the novel coronavirus pathogen. In Figure 1, we observe that the epidemic was controlled by the end of February, with almost no new cases. After this point, an increase in the number of recoveries is seen in Hubei. Our analysis particularly focuses on the historical data for the epicentre, Hubei province. The raw data contains the number of cumulative confirmed cases, cumulative recovered cases and cumulative deaths.

**Figure 1.**
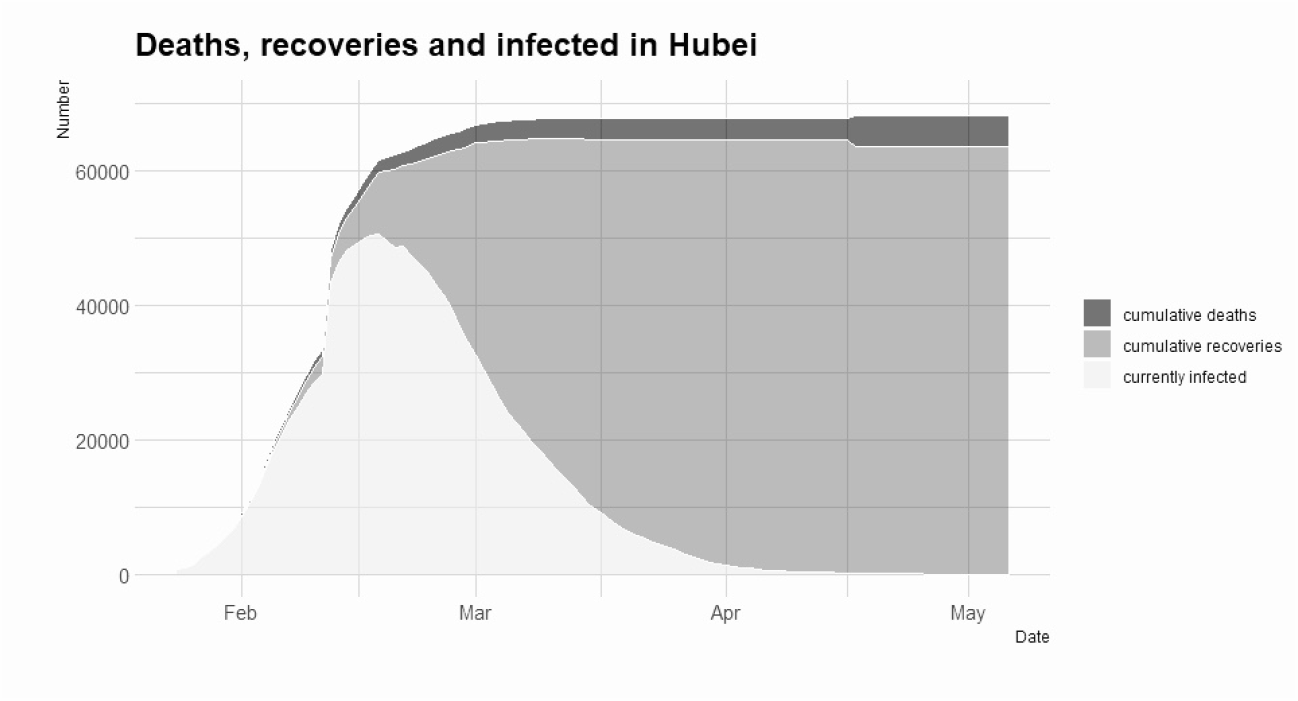
Number of cumulative infected, recovered and dead in Hubei

### 3.3 Descriptive analysis

Figure 2 shows the descriptive statistics for Hubei’s daily infection speed for the period 24 January - 17 March 2020, affected dramatically by the change in the way Chinese authorities accounted for confirmed cases as of 12 February. The mean and standard deviation graphs (Figures 2a and 2b) show sudden jumps on the day of the change, however values are fairly stable before and particularly after this point. Although the standard deviation experiences a slight increase and subsequent decrease over the period (without consideration of the 12 February peak), values remain below 0.005 throughout. The median (Figure 2a) is much more robust to the outliers in the data than the mean and is not largely affected by the significant jump in the number of cases. Dates highlighted in the plots represent elements of the timeline of particular significance in regard to the severity of protective measures. Irrespective of the jump on the change in definition, initially rising in value, the mean and median of the infection speed begin to fall on 5 February, around the time two specialist field hospitals were opened in the region. This decrease also occurs approximately one week after quarantine restrictions governed movement across all of Hubei and two weeks after the imposition of lockdown restrictions in Wuhan, displaying the effectiveness of lockdown measures with respect to infection speed. Two weeks after the opening of the first speciality hospital the infection speed begins a sharp decrease, before becoming more stable and ultimately reaching zero. This decrease is reflected in the standard deviation plot. Skewness and kurtosis values (Figures 2c and 2d) display similar patterns on different scales, both are mostly positive and consist of high values, an indication of a heavy tailed and right skewed distribution. There are no recorded deaths in Wuhan on 21 February which is reflected in the drop in skewness and kurtosis values.

**Figure 2.**
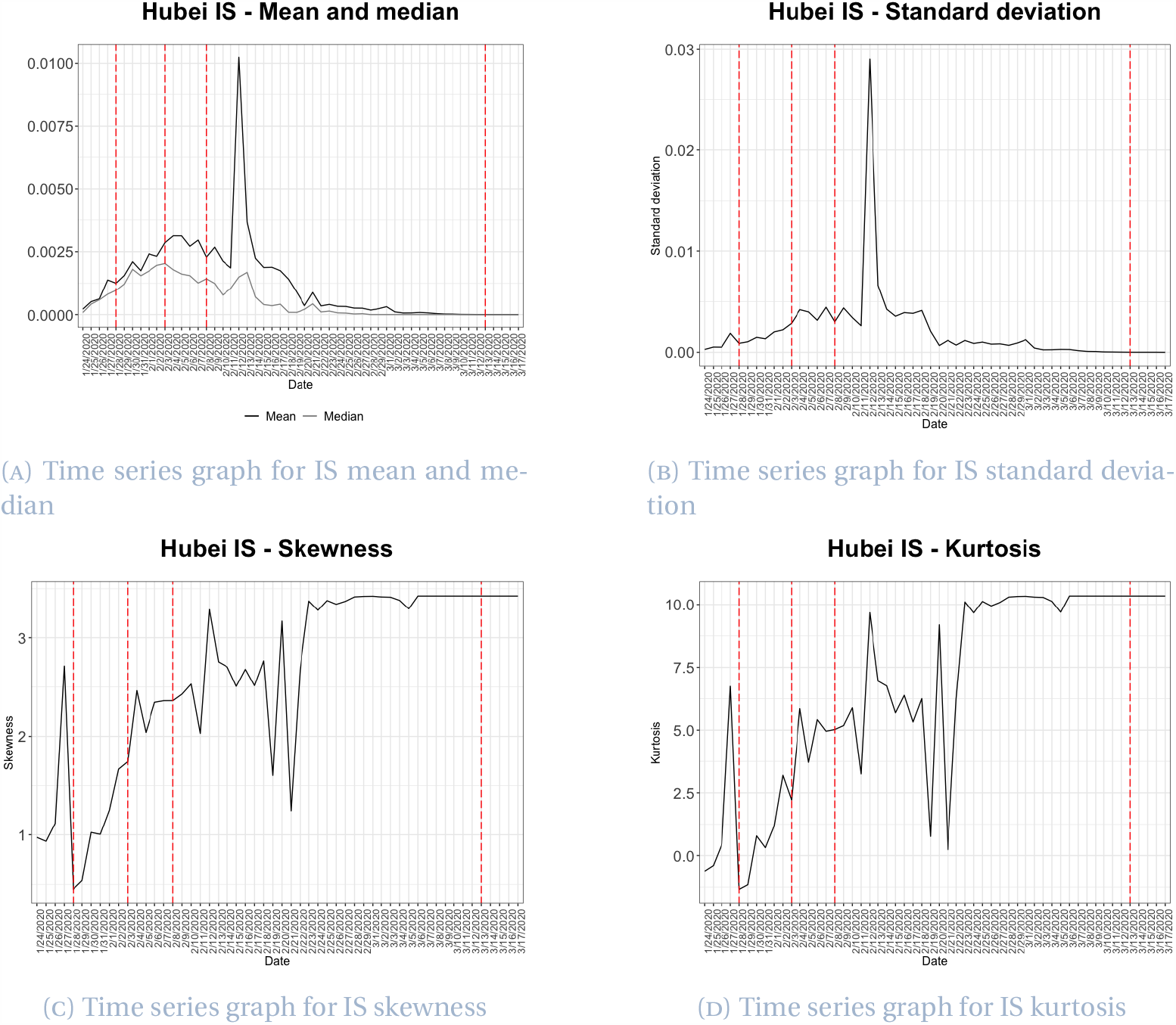
Descriptive statistics for Hubei’s infection speed

Displayed in Figure 3a, the mean and median daily death rate, investigated between 28 January and 17 March 2020, are fairly stable throughout the centre of the observed period and present a similar value trend from 6 February. Median values are however systematically lower than mean values, which may indicate a slight skewness to the right. Where the daily mean appears more stable between 14 February and 3 March, its average value is 0.0013, with both mean and median dropping to zero on 21 February as a result of the recording of no deaths on that day, almost two weeks after the opening of the second speciality hospital. The obvious downward trend in the mean plot after complete lockdown on 28 January shows the instant impact of the government reaction to the outbreak. Figure 3b presents a low and stable standard deviation, largely ranging from 0 to 0.005. Towards the end of the period, the mean and standard deviation of the death rate fluctuate slightly in the upwards direction. However, on all but two dates after 24 February (4 weeks after complete lockdown in Hubei), the median is zero. On 11 March, we observe a spike in the two values. Only 4 (including Wuhan) of the 17 cities in Hubei had a non-zero death rate on this day, suggesting the outbreak was under control in most cities at this point. Following the beginning of the easing of restrictions in Hubei, a significant spike is seen in Figures 3a and 3b on the final date of the observation period. The spike however, corresponds to a single death in Xianning, causing a deviation in their death rate from zero. The reduced death rate present throughout the majority of the period mainly corresponds to the decrease in the number of new infected cases and thus a decrease in the number of deaths in the region. Skewness (Figure 3c) mainly fluctuates between 0 and 3 despite taking a number of values both above and below this interval, whilst excess kurtosis (Figure 3d) is again extremely volatile and always positive, indicating a heavy tailed distribution for death rate time series data.

**Figure 3.**
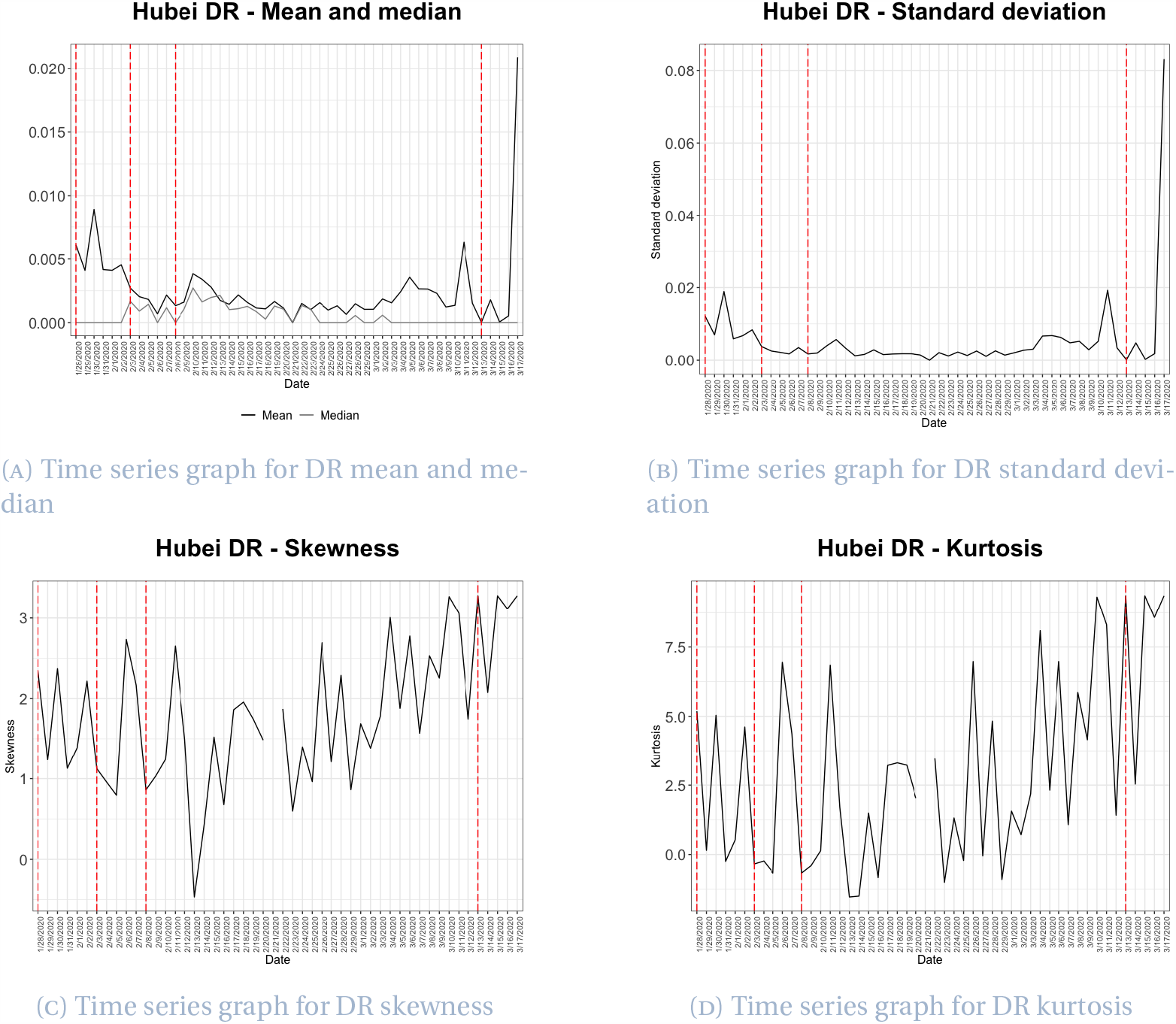
Descriptive statistics for Hubei’s death rate

Figure 4 presents graphs of the descriptive statistics for daily fatality ratios for the period 25 January - 17 March 2020. As seen in Figure 4a, the mean and median of daily fatality ratios are distant from one another during the early period of the outbreak, due to the recording of zero deaths in many cities. Mean and median values become closer around the middle of February, indicating the existence of a lack of outliers in the period under consideration, whilst also suggesting the distributions of daily fatality ratios are not skewed significantly. The two statistics follow the same trend after their intersection on 16 February, almost three weeks after lockdown restrictions governed all of Hubei. The average fatality rate falls slightly on the opening of the first speciality hospital on 3 March, however gradually increases around the time of the second hospital opening. During the latter half of the period of interest the mean stabilises at approximately 3%. In Figure 4b, we observe that the values of daily standard deviations are initially high, whilst concentrated at around 0.013 for the remainder of the period. The standard deviation of the fatality ratio experiences a significant decrease following its peak on 26 January, which appears to stabilise alongside the opening of the second speciality hospital. The decrease falls in line with introduction of lockdown measures across the whole of Hubei on 28 January, with the minimum variation in the data observed on 15 February, almost three weeks after lockdown was imposed. Taking positive values before 15 February and negative values afterwards, the skewness of the fatality ratio (Figure 4c) indicates that the data has different distributional structures, although skewness values are mostly between −1 and 1. Daily excess kurtosis values (Figure 4d) are more volatile and experience a similar change around 15 February, presenting greater absolute values before this point and relatively small positive and negative values afterwards. This change in distributional structure indicated in Figures 4a, 4c and 4d occurs in the third week of complete lockdown, perhaps reflecting the time at which mitigation measures substantially affected the fatality ratio in Hubei.

**Figure 4.**
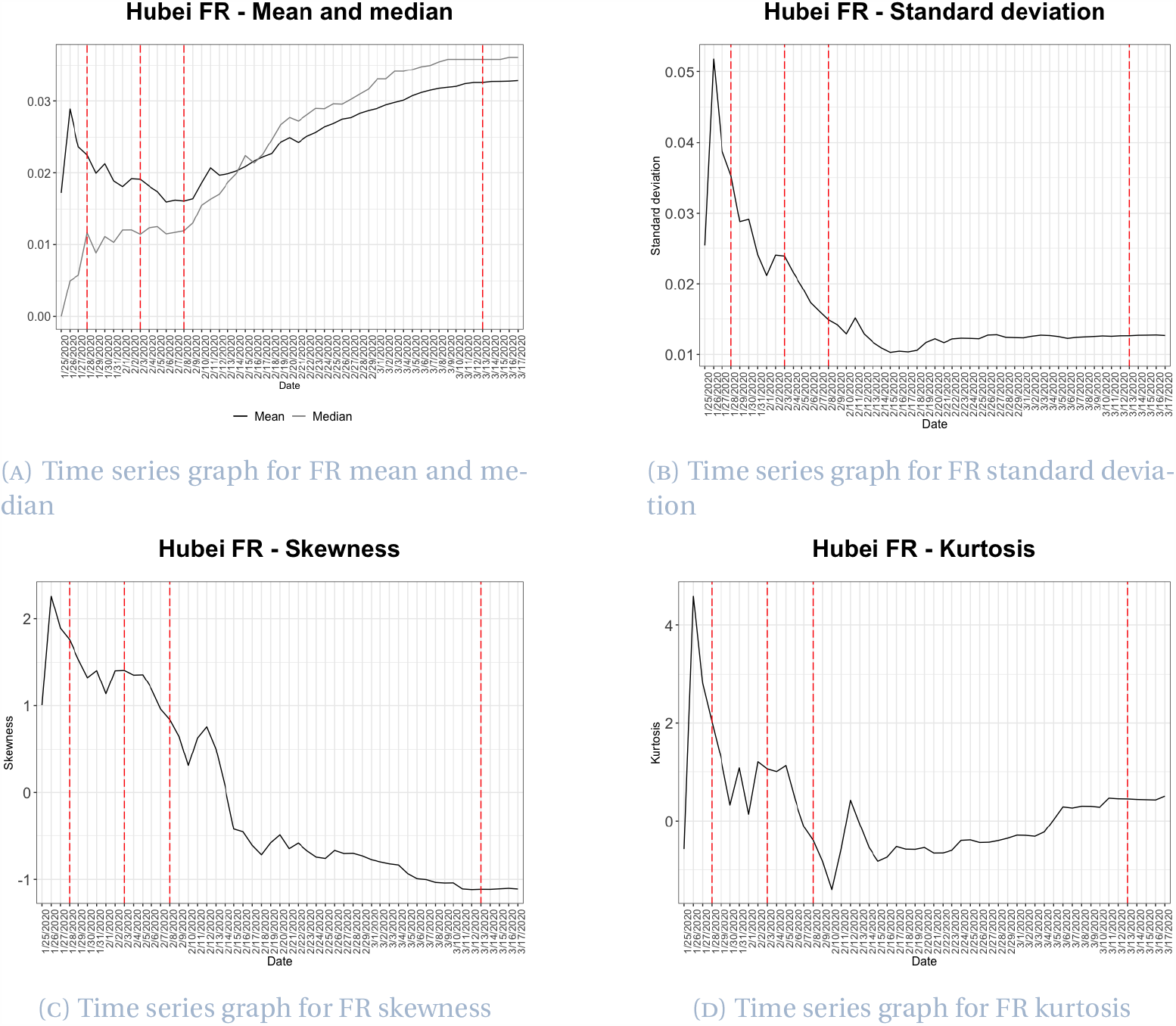
Descriptive statistics for Hubei’s fatality ratio

### 3.4 Distribution fitting

#### Infection speed

We consider infection speed as it reflects, in particular, government action in regard to the prevention of any further outbreak of the infection, through for example, isolation measures, travel restrictions and the provision of face masks. As discussed in Section 2.2, 61 distributions were fitted throughout period 24 January to 5 March. Observing the Kolmogorov-Smirnov test statistic for distributions fitted each day, we select the generalised extreme value (GEV) distribution as the fitting distribution for Hubei infection speed, and thus fit the GEV to all days in the observation period. The probability density function of the GEV distribution is given by

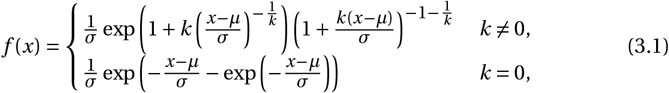

where *k* is the shape parameter, *σ* the scale parameter (*σ >* 0) and *µ* the location parameter.

GEV parameters for infection speed are plotted in Figure 5. The shape parameter *k*, displayed in Figure 5a, fluctuates throughout the period with an overall increasing trend between 0 and 1, stabilising slightly below 1 around 27 February, approximately one month after lockdown restrictions governed all of Hubei. The volatility of the daily *k* values is more significant during the first half of the period, with the reduction in the number of jumps with time suggesting some stabilising of the data. Significant jumps in the value of *k* occur on 2, 12 and 20 February. The positive jump on 12 February coincides with the change in the definition of confirmed cases, as in the mean and standard deviation descriptive statistics plots (Figures 2a and 2b). Daily parameters *µ* and *σ* (Figures 5b and 5c) both experience a sharp increasing and decreasing trend, before stabilising around 21 February. This increase and decrease is also reflected in the descriptive statistics plots for infection speed (Figure 2). Although both parameters behave in a volatile fashion, daily *µ* and *σ* parameters begin to decrease around 1 and 2 February, respectively, 4 and 5 days after lockdown restrictions were in place across the whole of Hubei. This decrease occurs almost simultaneously to the decrease in mean infection speed, displayed in Figure 2a. The change also coincides with the 2 February peak in the daily *k* values. On 18 February the minimum and only negative value of *µ* occurs. Positive values of the shape parameter *k* in Figure 5a indicate a heavy tailed distribution for infection speed throughout the pandemic, while the rapid convergence of the location and scale parameters to zero during the second half of the period is consistent with the daily infection speed data, for which values become smaller and approach zero in most cities. All three GEV parameter plots are most stable during the period succeeding 21 February. The change in the stability of the plots at this point in time is also observed in each of the descriptive statistics time series. This therefore suggests that 24 days after quarantine restrictions governed Hubei in entirety, the outbreak was controlled.

**Figure 5.**
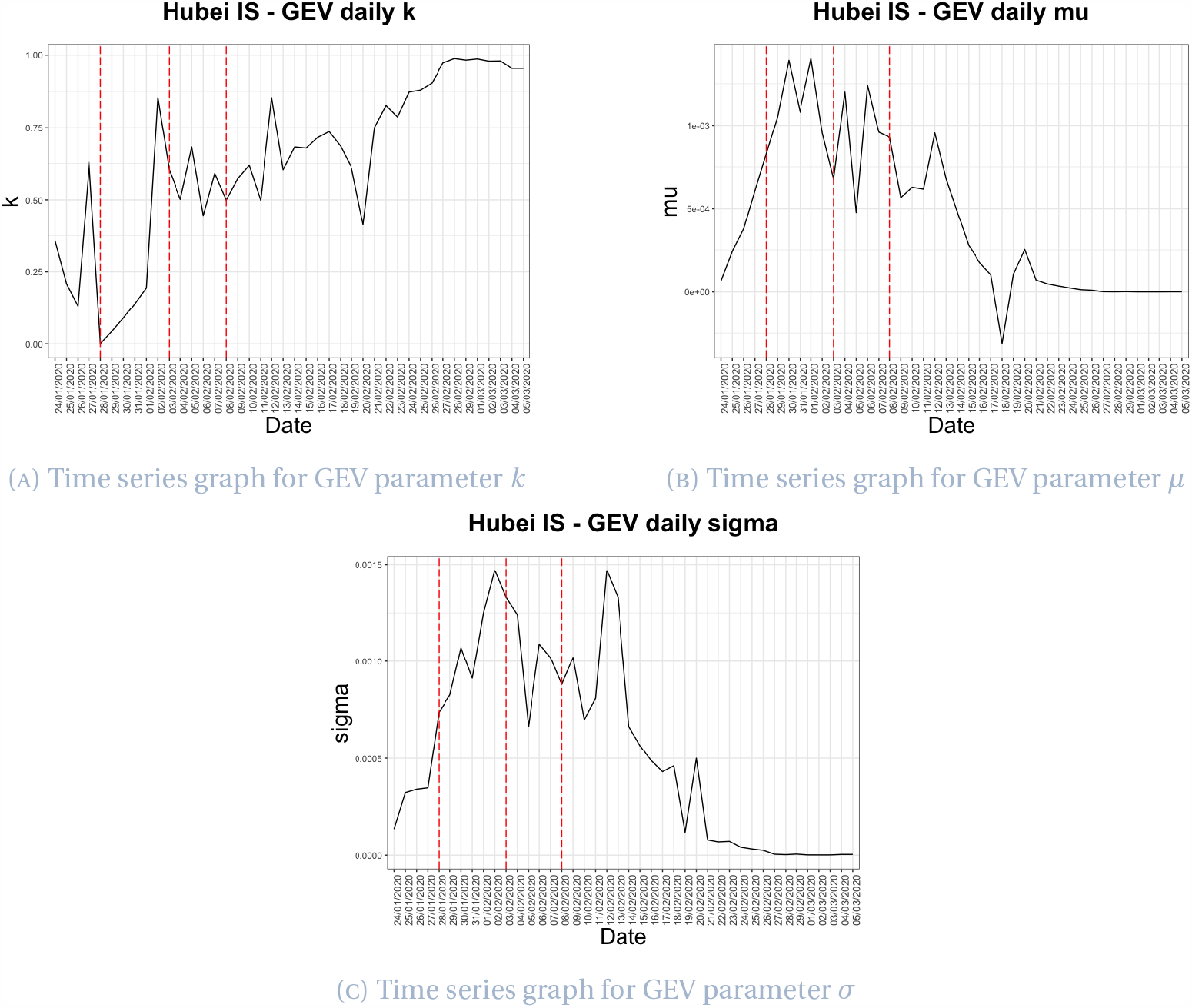
Time series for Hubei infection speed GEV parameters

#### Death rate

The Gumbel max distribution is the best fitting distribution for daily death rates, outperforming on 28 out of 49 days for the period 28 January to 16 March 2020.

The domain over which the distribution is defined is −*∞ < x < +∞* and the probability density function of the Gumbel max distribution is given by

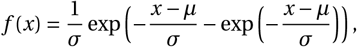

a special case of the GEV distribution when *k* is fixed at zero.

Daily values of the location parameter *µ* and scale parameter *σ* of the Gumbel max distribution are plotted in Figure 6. The parameter *µ* has a slight decreasing trend within the period, experiencing a number of spikes throughout. Values of *µ* are mostly positive however an increasing number of negative results occur within the final quarter. The minimum value of *µ*, recorded on 11 March, is −0.00236, significantly lower than all other parameter values. Three cities, in addition to Wuhan, have non-zero death rates on this date. Two spikes of almost equal height occur in the parameter *σ* both at the beginning and the end of the period. These spikes are also visible in the mean and standard deviation descriptive statistics plots (Figures 3a and 3b), with the later peak coinciding with the minimum value of *µ*. Apart from these two deviations, most values of *σ* fall between 0 and 0.005. A break in both plots occurs on 21 February due to the recording of no deaths. Considering Figure 6b, we observe an apparent stabilising of parameter values around the time of the opening of the speciality hospitals and 10 days after the introduction of region wide lockdown, at which time an immediate reduction in *σ* occurs. These trends are also displayed in descriptive statistics plots for death rate in Hubei (Figure 3) and may present evidence for the impact of government measures on reducing the death rate in the province. Note that the stabilising of the death rate appears to occur earlier than that of the infection speed, perhaps due to improvements in the availability of medical treatment and the experience of the medical profession in dealing with the outbreak over time. Values of *σ* become more volatile however towards the end of the period, just before the easing of restrictions.

**Figure 6.**
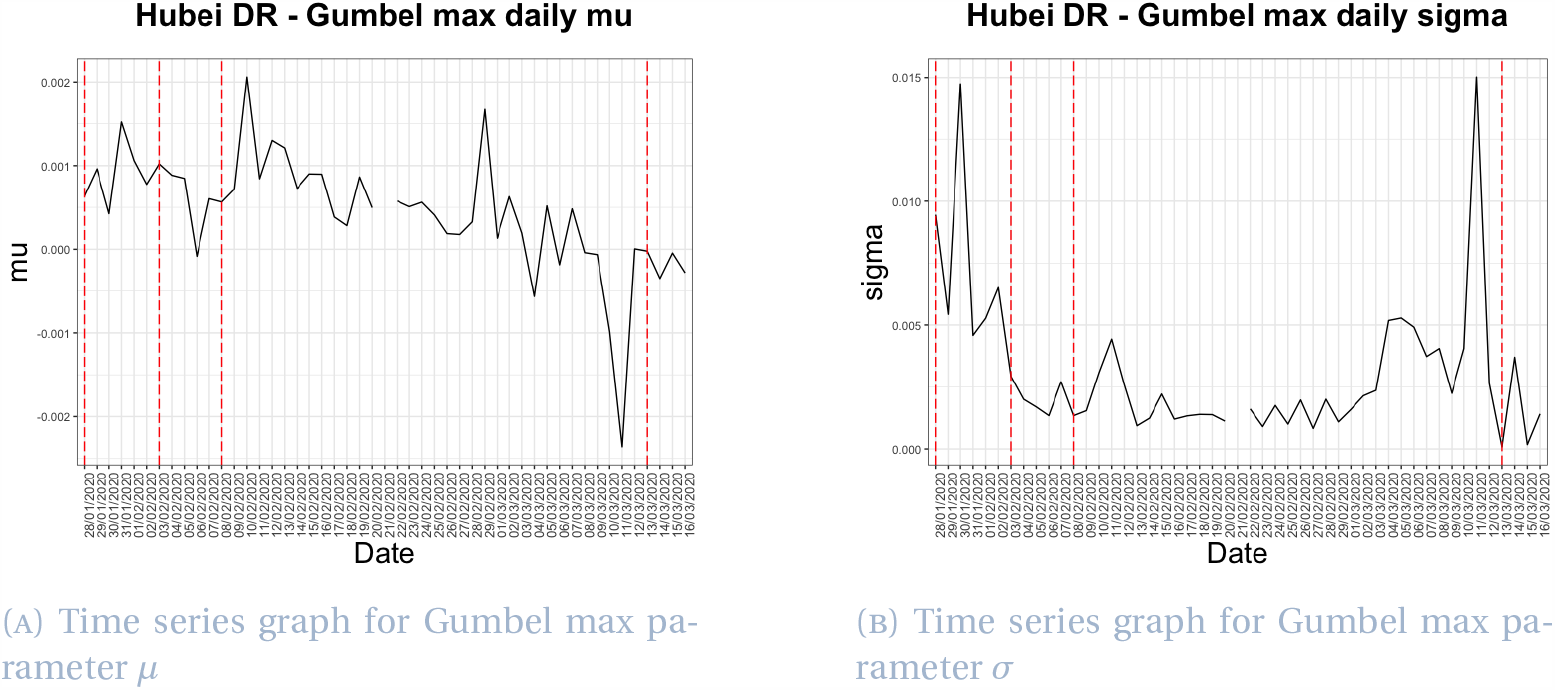
Time series for Hubei death rate Gumbel max parameters

#### Fatality ratio

For the final data set, we analyse daily fatality ratios for the 49-day period between 28 January and 16 March. Since fatality ratios are calculated through division of the cumulative number of deaths by the cumulative number of confirmed cases, they converge in line with the decreasing death rate and number of infected individuals. The GEV distribution is selected as the best fitting distribution in this case, outperforming on 28 days of the period under consideration. The probability density function of the GEV distribution is given in Equation (3.1).

Parameter values plotted in Figure 7 generally follow an increasing or decreasing trend throughout the observation period. Parameter *k* decreases from 0.54 to −0.89, whilst *µ* increases from 0.0050 to 0.032. Fluctuating more than parameters *k* and *µ*, the third parameter, *σ*, appears to stabilise around 22 February, in line with the distribution fitting for infection speed and two weeks after the opening of the second hospital. Occurring on 6 and 11 February, two *σ* spikes appear two weeks after the declaration of lockdowns in Wuhan and Hubei, respectively. However, the jumps move in opposite directions. Slight changes in the general trend across the period around 11 February are also seen in the descriptive statistics plots (Figure 4). Whilst the change in *µ* is fairly consistent throughout the period, experiencing a slight increase in gradient on 6 February followed by a decrease around 21 February, the rate of change of *k* is clearly split into two sections. Decreasing at a faster rate during the first three weeks of the period (and the first three weeks of lockdown), the trend in daily *k* values changes around 18 February, with the decrease in *k* becoming more stable. The change in all three parameters between 18 and 22 February (during the fourth week of regional lockdown) provides further evidence to support suggestion that a change in the severity of the outbreak occurred within this period, triggered by the government measures.

**Figure 7.**
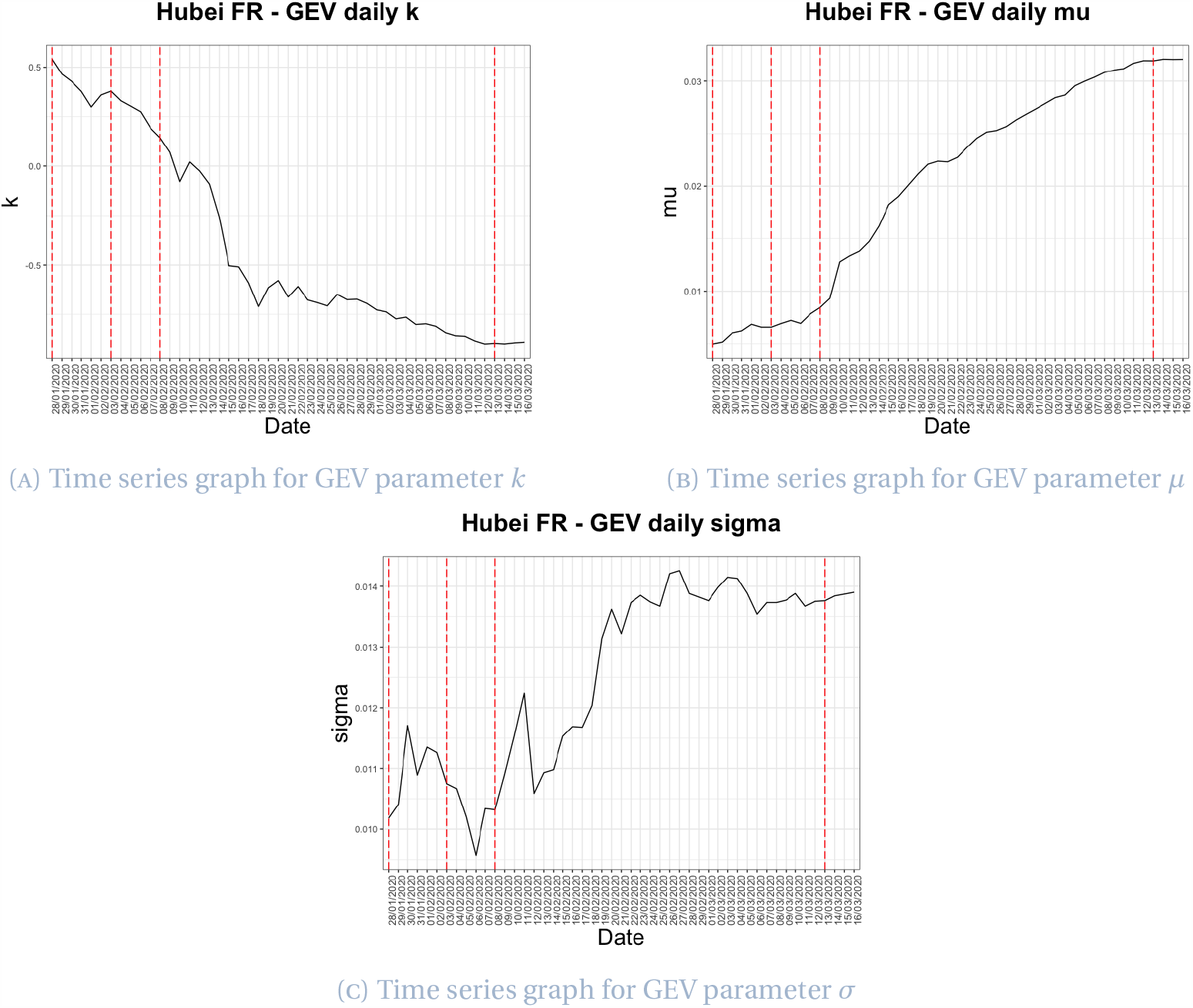
Time series for Hubei fatality ratio GEV parameters

## 4 Italy

### 4.1 Timeline of events

In this section, we present the timeline of COVID-19 events in Italy from 31 January 2020 to 25 May 2020.

#### First confirmed cases (31 January - 19 February)

On 31 January 2020, the first two cases of COVID-19 were confirmed in Rome ^23^. The Italian government suspended all flights to and from China and declared a state of emergency with a duration of six months the same day ^24^.

#### Clusters of cases in Northern Italy (20 February - 24 February)

The number of confirmed cases in Italy increased on 21 February, with sixteen people in Lombardy and Veneto confirmed to be infected ^25^. Following the first two deaths of individuals with the virus, around 22 February, ten municipalities of the province of Lodi in Lombardy and one in the province of Padua in Veneto were placed on lockdown due to the large number of infected cases in the region ^26 27^. Lockdown restrictions were initially implemented until 6 March. While residents were permitted to leave their homes, with supplies including food and medication allowed to enter, public gatherings and attending schools or workplaces were prohibited ^28^.

#### The spreading of the virus to other regions (25 February - 9 March)

After 25 February, cases emerged in several Italian regions without a clear link to the Northern Italy clusters. On 4 March, the Italian government imposed a nationwide two week shutdown of all schools and universities, as the country reached 100 deaths due to the outbreak ^29 30^. On the night of 7 March, Prime Minister Conte extended the lockdown to cover the whole region of Lombardy and 14 other northern provinces in Veneto, Emilia-Romagna, Piedmont and Marche, impacting more than 16 million people ^31^.

#### National lockdown (10 March - 3 May)

On 10 March, the Prime Minister again extended lockdown such that it governed all of Italy, implementing travel restrictions and a complete ban on public gatherings ^32^. All restaurants and bars were closed from 11 March, nationwide. On 21 March, further restrictions were announced within the nationwide lockdown, halting all nonessential production, industry and business ^33^. On 1 April, the government extended the period of lockdown to 13 April ^34^. On 8 April, all Italian ports were closed until 31 July as a result of a government decree ^35^. On 10 April, lockdown was prolonged until 3 May, as the reopening of a number of businesses including bookshops and foresters was permitted ^36 37^. Three weeks into lockdown, its impact began to show. On 20 April 2020, Italy saw its first fall in the number of active cases ^38^.

#### Easing of lockdown (4 May -)

On 26 April, the Prime Minister announced a starter plan for the so-called “phase 2”, that would begin on 4 May and allowed the re-opening of manufacturing industries and construction sites. From 18 May most businesses were able to reopen, and free movement was granted to all citizens within their region; movement across regions was still prohibited for non-essential reasons. On 25 May, swimming pools and gyms reopened, and on 15 June theatres and cinemas followed ^39^.

### 4.2 Data

The Italian Department of Civil Protection uploads daily Italian regional COVID-19 data on GitHub [7]. This data set includes daily information regarding the cumulative number of confirmed cases, cumulative deaths, cumulative recovered cases and newly infected cases, in addition to a number of other statistics which are not incorporated in this project. Data in the repository documents the information for each region in Italy from 24 February 2020 to date (at the time of writing).

In Figure 8, we observe the slowing down of the epidemic around the middle of April, where the number of active cases reaches its peak, potentially as a result of lockdown measures. For the remainder of the observation period, an increase in the number of recoveries is observed.

**Figure 8.**
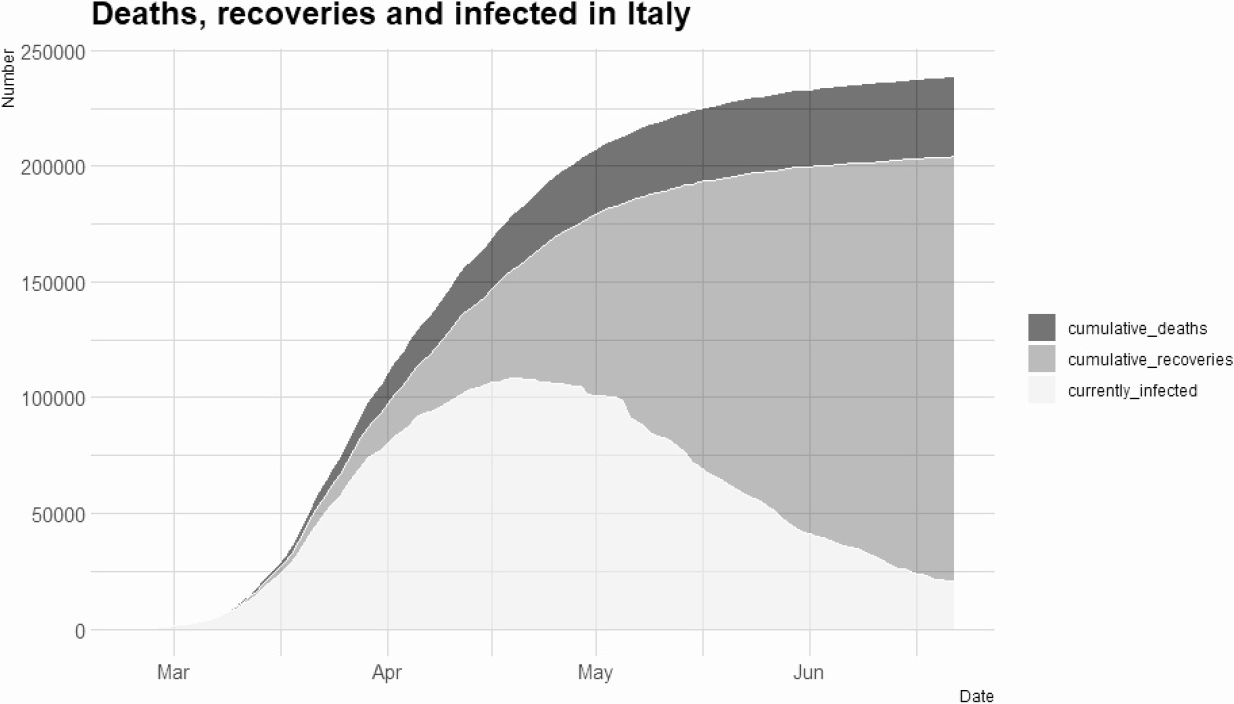
Number of cumulative infected, recovered and dead in Italy

### 4.3 Descriptive analysis

Descriptive statistics for Italian infection speed data are displayed in Figure 9 for the period 25 February – 15 June 2020. Again, the skewness (Figure 9c) and kurtosis (Figure 9d) follow similar patterns but on different scales with their peaks coinciding. Almost all values of skewness and kurtosis are positive and greater than 1 for the duration of the period, supporting selection of a heavy tailed distribution for infection speed. The mean infection speed (Figure 9a) peaks on 28 March, just over two weeks after the beginning of the nationwide lockdown and one week after the halting of non-essential business, decreasing in general after this point. The subsequent decreasing trend provides indication of the effectiveness of the lockdown measures in reducing the infection speed. Between 15 March and 20 April, the mean infection speed is highly volatile, with the rate of change of infection speed greatest between 10 March and 21 March. The median (Figure 9a) follows a similar pattern to the mean however is generally lower and more volatile, indicating a slight right skewness in the daily infection speed distributions. The standard deviation of infection speed (Figure 9b) is volatile during the same period before peaking on 18 April, at which time the occurrence of a spike is observed in the mean but with less significance. The measure stabilises towards the end of the period in line with both the mean and the median and the easing of the outbreak.

**Figure 9.**
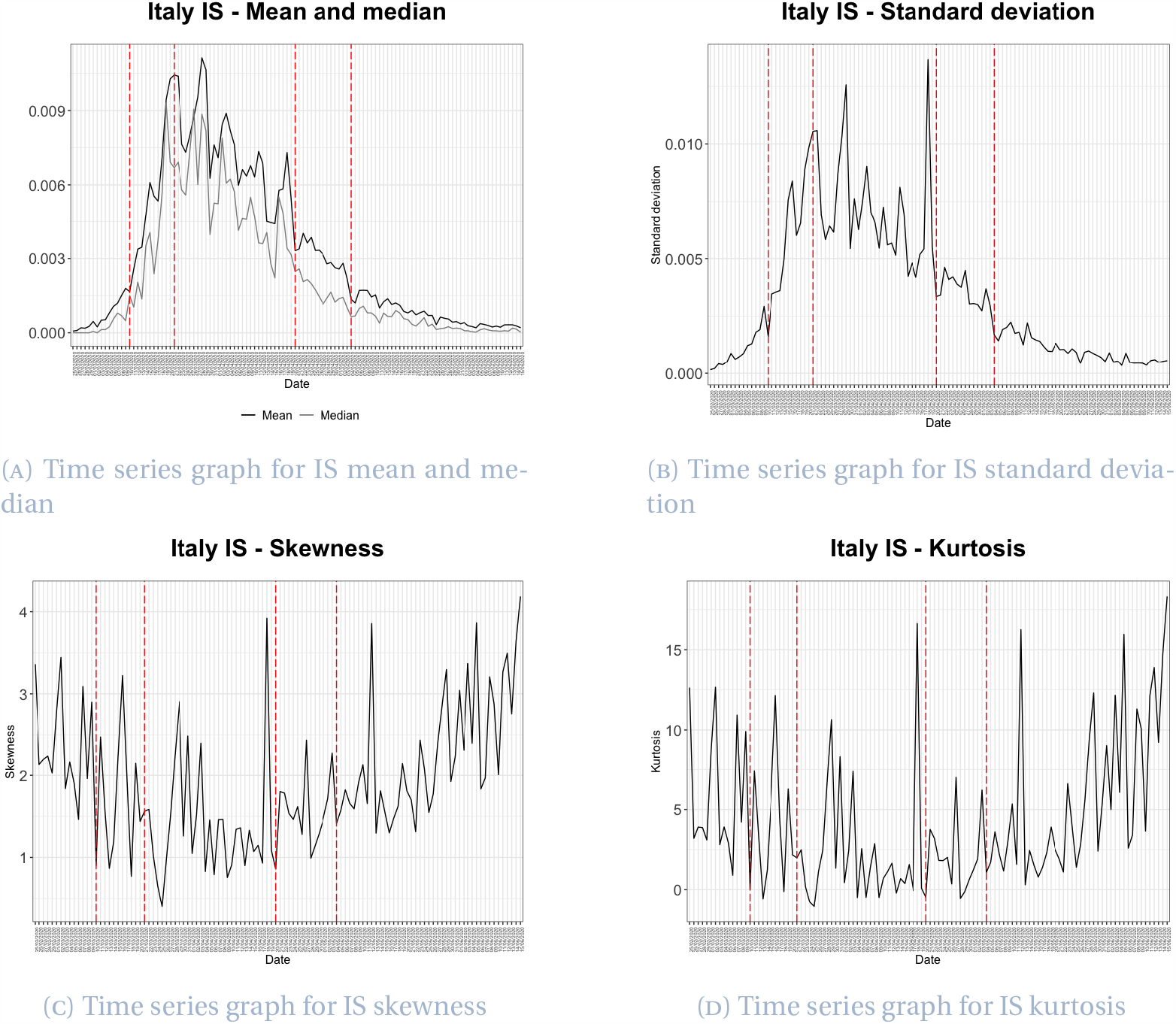
Descriptive statistics for Italian infection speed

Descriptive statistics for Italian death rate data for the period 25 February – 15 June 2020 are displayed in Figure 10. Considering first the mean and median of the data in Figure 10a, the mean death rate peaks on 19 March, around two weeks after the closing of all schools and universities across the country on 4 March, the date of the first significant jump in death rate. The mean data also experiences a later peak on 6 June, coinciding with the easing of the lockdown restrictions. The first fall in the number of cases was recorded on 20 April however by this point the mean death rate is more controlled and decreasing with time. Values of the mean and median differ initially due to the recording of zero deaths in many regions at the beginning of the outbreak. After the median jump on 12 March, the mean and median coincide, indicating that daily death rate distributions are not significantly skewed. Considering the standard deviation of the death rate displayed in Figure 10b, the most significant peak occurs on 4 March, with a second peak on 6 June. The standard deviation is less volatile than the mean and appears fairly stable between 7 April and 28 May, fluctuating around 0.0025. Although kurtosis (Figure 10d) is the more volatile measure for daily death rates, skewness (Figure 10c) and kurtosis behave similarly across the period with differing scales, experiencing an initial decrease before rising around the time of the first drop in the number of cases. Both skewness and kurtosis are mostly positive, with the majority of skewness values fluctuating between 0 and 3 and only 6 skewness values falling below zero across the whole period. The high values of skewness and kurtosis indicate the appropriateness of a heavy tailed distribution for Italian death rate data.

**Figure 10.**
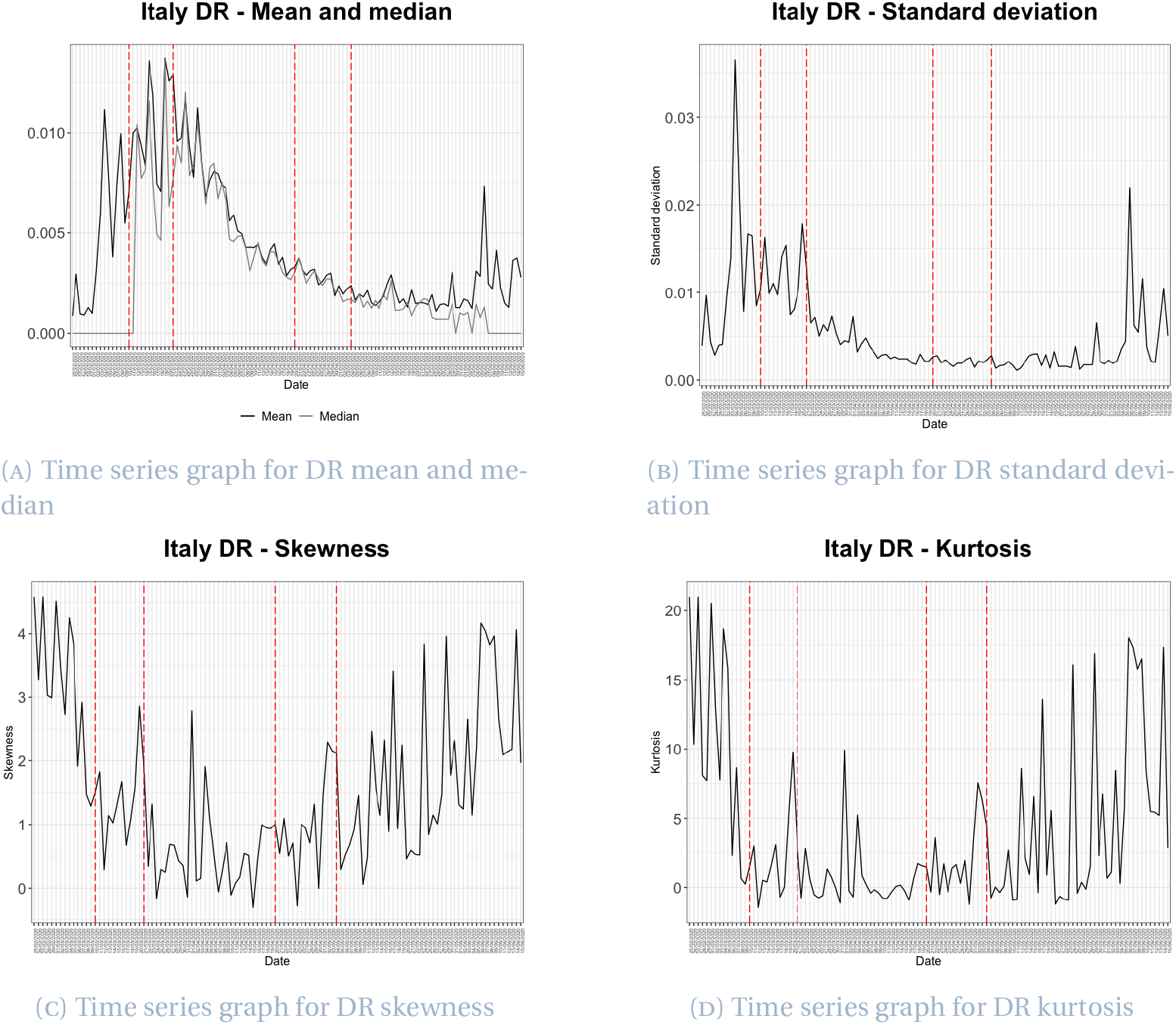
Descriptive statistics for Italian death rate

Figure 11 shows the descriptive statistics for Italian fatality ratio for the period 25 February – 15 June 2020. The mean fatality ratio (Figure 11a) increases steadily throughout. A slight bump in the data occurs before the first lockdown on 7 March, after which the mean increases steeply until around 7 April, almost one month after the declaration of nationwide lockdown, when the rate of increase begins to slow. Both mean and median (Figure 11a) remain fairly steady after this point. Following a slight jump on 11 March after exiting the period of mostly zero death recordings, the median follows the same pattern as the mean however is generally lower, suggesting the data is slightly skewed to the right. The standard deviation of the fatality ratio (Figure 11b) also appears to stabilise around 12 April, becoming steady around a value of 0.03 in the latter third of the period as the virus starts to become more under control and restrictions are eased. The first jump in the standard deviation occurs on 5 March and the peak is on 9 April, with the measure behaving volatilely between these points. Figures 11c and 11d highlight the stabilising of skewness and kurtosis over time although initially volatile, particularly in the kurtosis case. While after 8 March kurtosis largely fluctuates between −1 and 1, between 8 March and 25 March, kurtosis values are mostly negative. Data for all other dates before 8 June are associated with positive kurtosis. Skewness is positive throughout the period, remaining below one in most cases after 9 March. The change in skewness and kurtosis values around this point in time and their continued decrease suggests the structure of distributions for the daily data differs within the period, moving from highly skewed and heavy tailed to almost symmetric. The fatality rate data for each region stabilises around 20 May, at which point the lockdown measures had eased significantly. This cannot be observed in the death rate and infection speed data.

**Figure 11.**
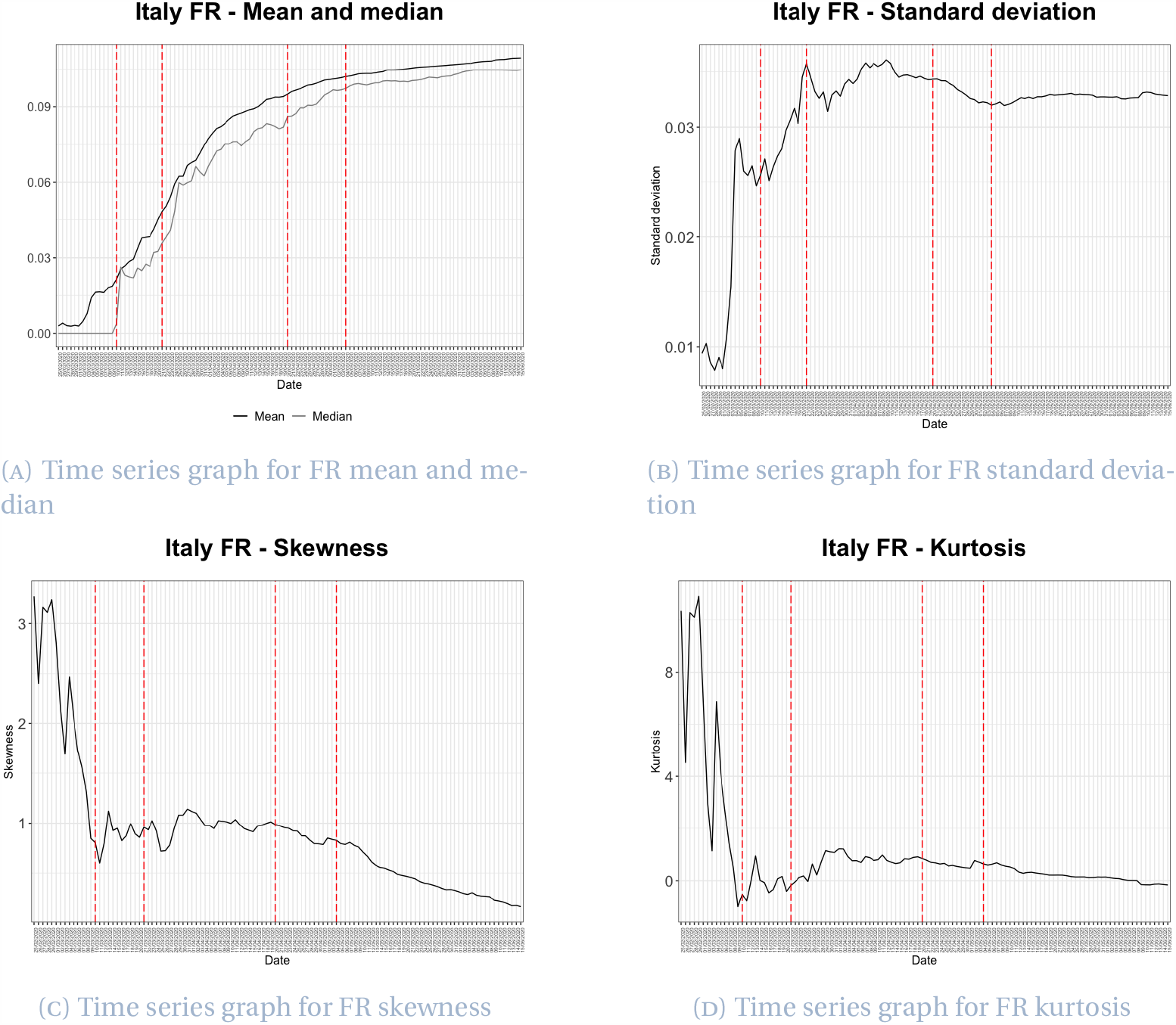
Descriptive statistics for Italian fatality ratio

### 4.4 Distribution fitting

#### Infection speed

After fitting various distributions for the period 25 February - 15 June, we select the generalised Pareto distribution as the best fitting distribution for Italian infection speed data, outperforming on 74 days of the 112 day period. The probability density function of the generalised Pareto distribution is given by

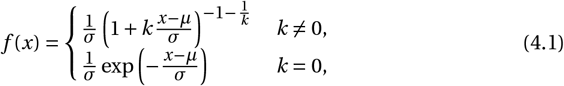

where *k* is the shape parameter, *σ* the scale parameter (*σ >* 0) and *µ* the location parameter. The domain over which the distribution is defined is

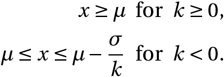

Time series plots of infection speed parameters are shown in Figure 12. The parameter *k* is the most volatile throughout the period, varying from −0.52 to 0.81. However, the majority of the time series plot lies in the positive portion of the graph. Approximately two weeks after the declaration of nationwide lockdown on 10 March, daily *k* values begin to increase, after decreasing throughout the initial part of the observation period. Almost three weeks after the first easing of restrictions on 4 May, *k* experiences a negative spike, although the overall trend continues to increase. The parameter *µ* starts to fluctuate in line with the beginning of nationwide lockdown and stabilises as lockdown is lifted on 3 May, behaving as a process mean-reverting about zero and peaking on 28 March. The third parameter of the generalised Pareto distribution, *σ*, follows a similar pattern to both *k* and *µ*, volatile at the beginning of the lockdown and experiencing a sharp increase, with peak on 25 March, two weeks after the declaration, before falling after this point with decreasing volatility. The consistent pattern in each of the three plots suggests lockdown measures in Italy took around two weeks for an obvious impact on infection speed to be observed. The clarity of the period of highest severity in the three parameter plots (particularly *µ* and *σ*) is parallel to that of the mean, median and standard deviation descriptive statistics plots in Figures 9a and 9b. This highlights the significance of government measures in the controlling of infection speed in Italy.

**Figure 12.**
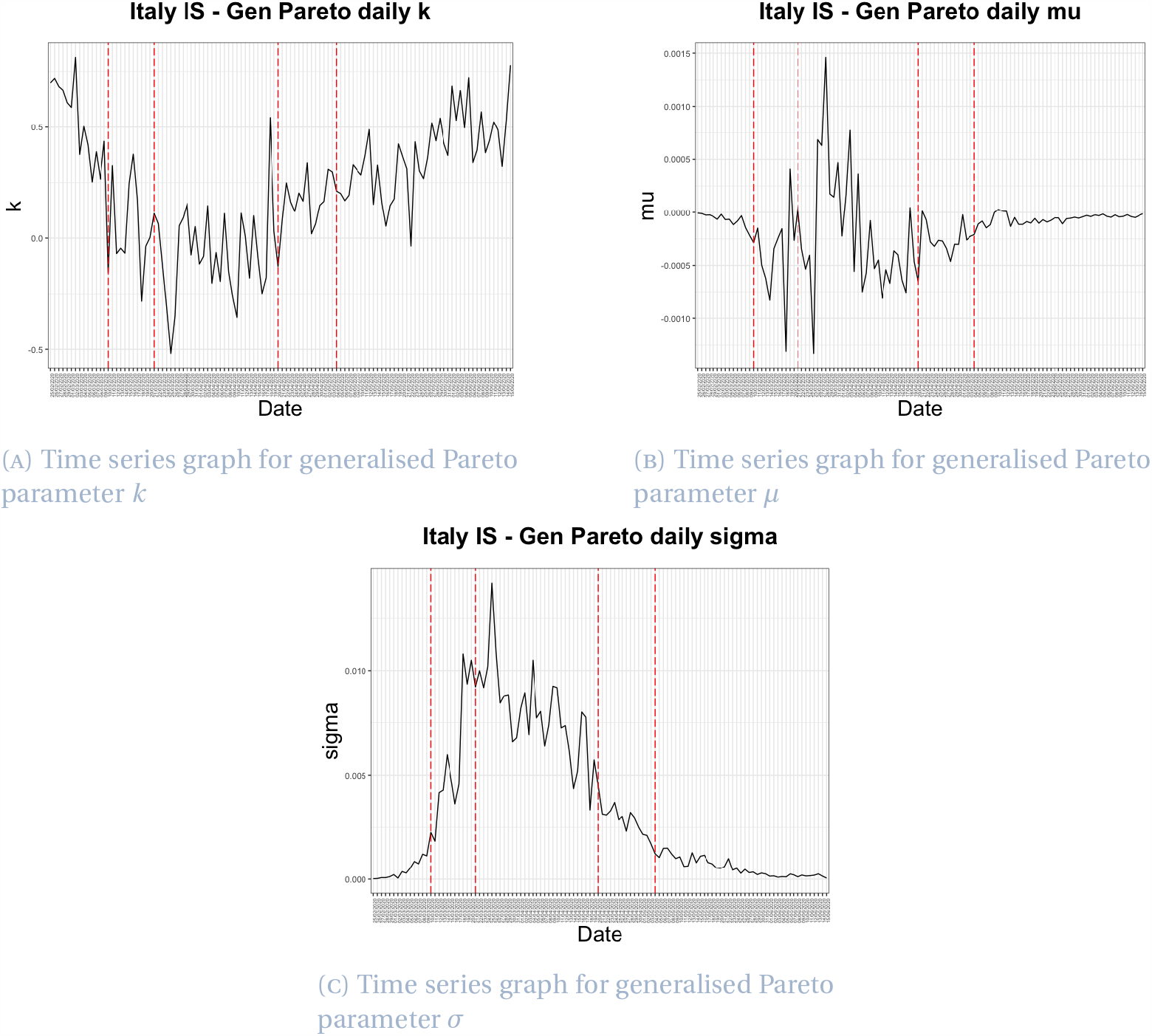
Time series for Italy infection speed generalised Pareto parameters

#### Death rate

The Johnson S_B_ (JSB) distribution outperforms for death rate on 46 days of the 105 day period between 3 March and 15 June, and is therefore chosen as the best fitting distribution. The density function is given by

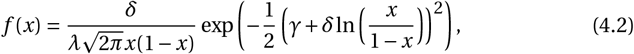

where *γ* and *δ* are shape parameters (*δ >* 0), *λ* is the scale parameter (*λ >* 0) and *ξ* the location parameter. The domain over which the distribution is defined is *ξ ≤ x ≤ ξ+λ*. Plots of the four parameters of the JSB distribution are shown in Figure 13. In general, all parameters are volatile throughout the whole period. The daily *γ* values largely fluctuate between 0 and 2, with a sudden spike of 11.29 occurring on 19 April, the day before the first fall in the number of active cases was confirmed and almost six weeks after the beginning of nationwide lockdown. The parameter *δ* moves between 0 and 2.5 with the exception of 19 April, in line with the *γ* peak. The *δ* parameter values, although extremely volatile, appear to behave in a gradually increasing then decreasing fashion. The maximum daily *λ* value also occurs on 19 April, largely remaining between 0 and 0.05 after this point. Generally fluctuating within a small interval, the parameter *ξ* has a range of 0.012. The volatility in this parameter is particularly large between 10 March and 4 May, the period during which lockdown restrictions were most severe. We observe the greatest volatility in *ξ* at the centre of this lockdown period, with extremely volatile rates occurring towards the end of March, three weeks after the imposition of lockdown and around the time of its first extension. This volatility suggests that at the point of its extension, the outbreak in Italy was not under control. The stabilising of all parameters towards the end of this period may display the effectiveness of the prolonged sustaining of restriction measures.

**Figure 13.**
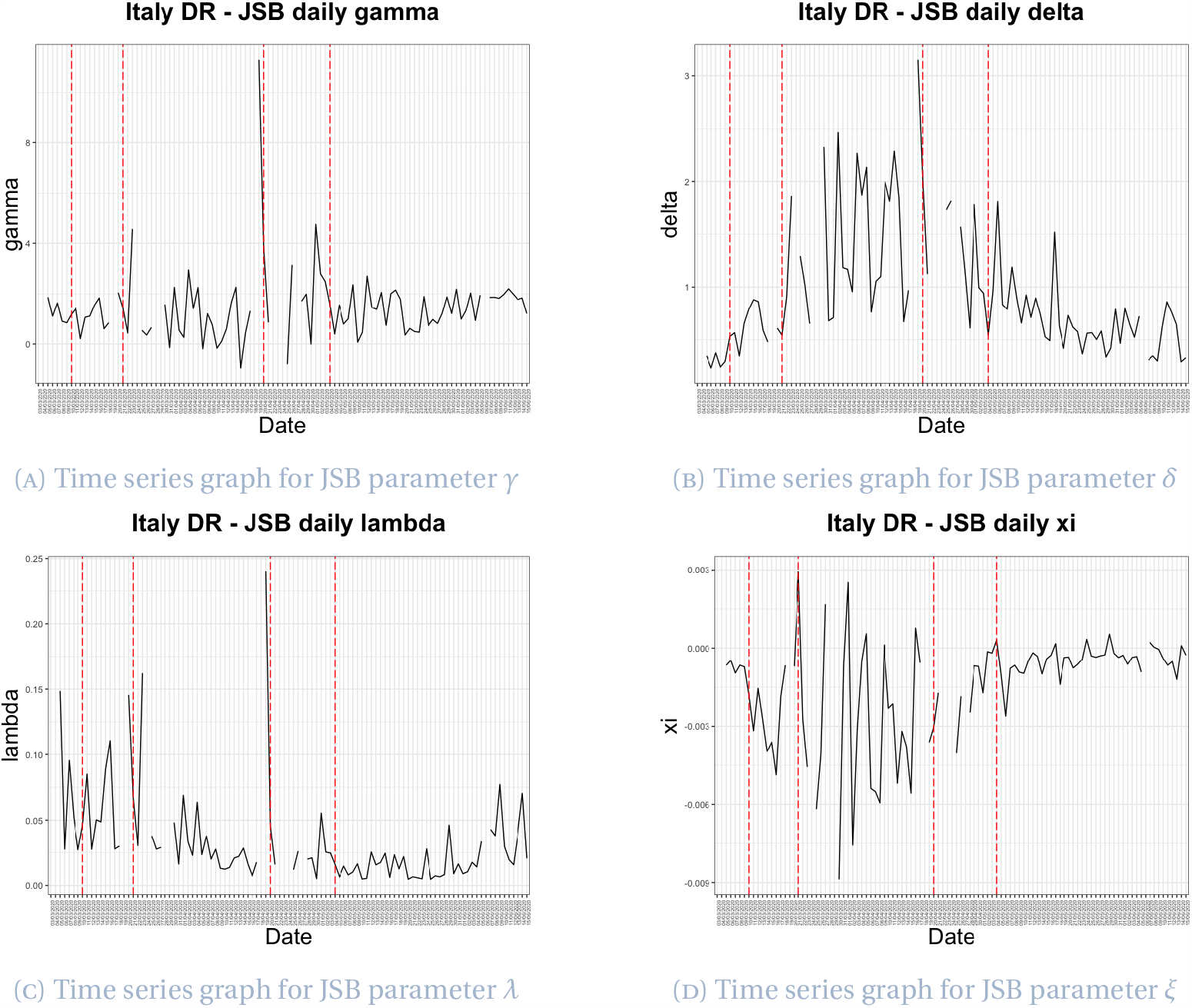
Time series for Italy death rate Johnson S_B_ parameters

All four plots in Figure 13 include a number of breaks, these breaks correspond to dates for which the JSB distribution provided no fit to the data. Of particular significance are the consecutive breaks between 18 April and 27 April, with just 4 dates fitting to the JSB distribution within this period. This fitting failure occurs around the time the first fall in cases was observed, implying there was a clear change in the death rate at this point of the outbreak in Italy. This cannot be observed when simply considering the associated descriptive statistics plots in Figure 10.

#### Fatality ratio

The Johnson S_B_ distribution again dominates for Italian fatality ratio data, as it outperforms on 33 days out of the 105 day period between 3 March and 15 June.

Plots in Figure 14 display the time series for all four JSB parameters throughout the investigation period. We observe that parameters *γ, δ* and *λ* have similar trajectories, with their peaks occurring simultaneously on 1 June, at which time Italy was within phases 2 and 3 of the easing of lockdown restrictions. The trajectory for *ξ* however, is a reflection of the other three parameters, with its minimum occurring on 1 June. There therefore appears to be a definite change in the fatality ratio data on this date, with the reversal of the general trend in all four cases. Whilst *γ, δ* and *λ* are mostly positive, *ξ* is mostly negative. A spike in all four plots occurs on 26 March, approximately two weeks after the declaration of nationwide lockdown. Change on this date is observed only slightly in the descriptive statistics plots (Figure 11), with no noteworthy peak. All plots in Figure 14 stabilise around 1 April, the day of the lockdown extension, until approximately 23 April, at which point the trend changes, with a sharp increasing (or decreasing in the *ξ* case) trend, before reaching the 1 June peak. Although a clear change in each of the parameter plots is seen both leading up to 1 June and after this point, the descriptive statistics plots in Figure 11 experience steady trends around this time in the observation period.

**Figure 14.**
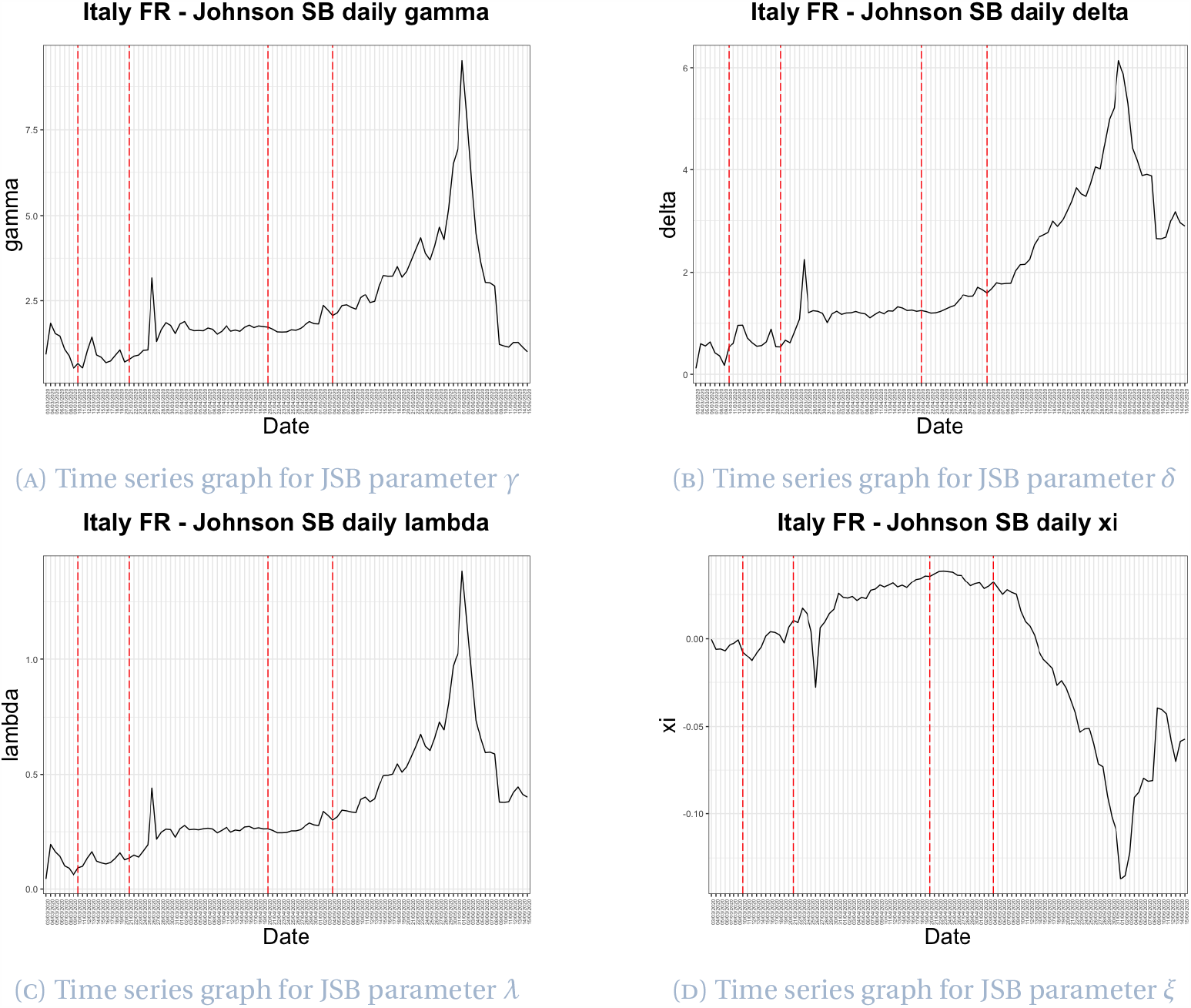
Time series for Italy fatality ratio Johnson S_B_ parameters

## 5 Spain

### 5.1 Timeline of events

We now present the timeline of COVID-19 events in Spain from 31 January 2020 to 21 June 2020.

#### First confirmed cases (31 January – 25 February)

The pandemic was first officially confirmed to have spread to Spain on 31 January, when a German tourist tested positive for SARS-CoV-2 in La Gomera, Canary Islands, having been in contact with individuals who had travelled to China ^40 41^. On 13 February, the death of a 69-year-old man who had travelled from Nepal was recorded as the first death.

#### Community transmission (26 February – 12 March)

On 26 February, the first reported case in the region of Andalusia was confirmed in Seville, becoming the first recognised case of community transmission in Spain. On 7 March, Haro was the first town to be put on lockdown, due to a high concentration of cases ^42^. On 10 March the Government of Spain decreed the immediate cancellation of all direct flights from Italy to Spain until 25 March ^43^.

#### State of emergency (13 March – 27 March)

On 13 March, the Prime Minister of Spain declared a 15 day nationwide state of alarm (the lowest level of a state of emergency), to become effective the following day after approval by the Council of Ministers ^44^. On 14 March, the Spanish government imposed the nationwide lockdown, banning all trips that did not qualify as force majeure ^45 46^. A state of emergency was declared the next day and the imposition of a national lockdown beginning 15 March announced as part of emergency measures to combat the coronavirus outbreak in the country ^47 48^. Individuals were permitted to leave their homes only for valid reasons, including shopping, visiting vulnerable relatives, attending medical appointments and travelling to work for key workers only. On 16 March, the Minister of the Interior Grande-Marlaska announced the closing of Spanish frontiers as of 12:00 pm on 16 March, authorising the entry of only Spanish citizens and those who could prove cause of force majeure or a situation of need.

#### Halting of all non-essential activity and initial extension of state of alarm (28 March – 12 April)

On 28 March, the Spanish government banned all non-essential activities ^49 50^. On 4 April, Prime Minister Pedro Sánchez asked the Congress of Deputies to extend the state of emergency for another two weeks, until 26 April, a request that was granted on 9 April ^51^.

#### Easing of restrictions and second extension of state of alarm (13 April – 1 May)

On 13 April, workers in a number of non-essential sectors, including the construction and industry sectors, who could not work remotely were permitted to return to work. On 23 April, the state of emergency was extended until 9 May, with further extensions envisioned ^52^.

#### De-escalation (2 May –)

On 28 April, the government announced a plan for easing lockdown restrictions which consisted of four phases. Phase 0 allowed citizens to leave their homes for short walks and individual sports from 2 May. Phase 1 was introduced on 11 May in 26 provinces and territories comprising about half of the Spanish population ^53^, and included the opening of small shops, in addition to terraces and places of worship, at half and one-third capacity, respectively. Phase 2 began on 18 May in a number of Balearic Islands; simultaneously, the remainder of Andalusia and the Valencian Community, alongside some territories in Castile and León and in Catalonia progressed to phase 1. As of 25 May, 47% of the territory of Spain was within phase 2 ^54^. As of 8 June, 48% of the country was within phase 2 and 52% within phase 3; the latter phase included less stringent restrictions for the opening of shops, hotels, bars, entertainment and nightlife venues. As of 15 June, 75% of the Spanish population was within phase 3 and several provinces entered the “new normality” phase ^55^. The state of emergency expired at midnight on Sunday 21 June, and the whole of Spain entered the “new normality” phase, in which restrictions such as maximum occupancy in shops was to be handled by each autonomous community independently ^56^. The government also opened all internal borders among autonomous communities as well as the land border with France, and resumed international flights with other European Union countries and the United Kingdom ^57^.

### 5.2 Data

Since 26 February 2020, the Coordination Center for Health Alerts and Emergencies (CCASE), a body of the Spanish Ministry of Health, has published a daily COVID-19 report for Spain [2]. The report contains daily information on the cumulative number of confirmed cases, cumulative recovered cases, cumulative deaths, and newly infected cases for each region in Spain from 25 February 2020 to 14 April 2020. After 15 April, the Coordination Center began to present the number of cumulative confirmed cases such that it consists of two parts, positive cases detected by a polymerase chain reaction (PCR) test and positive cases detected through an antibody test ^58^. Since the group of individuals with a positive antibody test may have been infected a number of days earlier, we decided to use the positive cases detected with PCR in order to represent the cumulative confirmed cases. The Coordination Center for Health Alerts and Emergencies no longer provided the number of recoveries as of 17 May. In the Spanish case, we consider only infection speed and fatality ratio due to the termination of death rate data announcements per region and hence the occurrence of missing regional data, with only complete national death rate data available.

Figure 15 shows the dynamic evolution of cumulative deaths, cumulative recoveries and active cases in Spain up to 14 April 2020.

**Figure 15.**
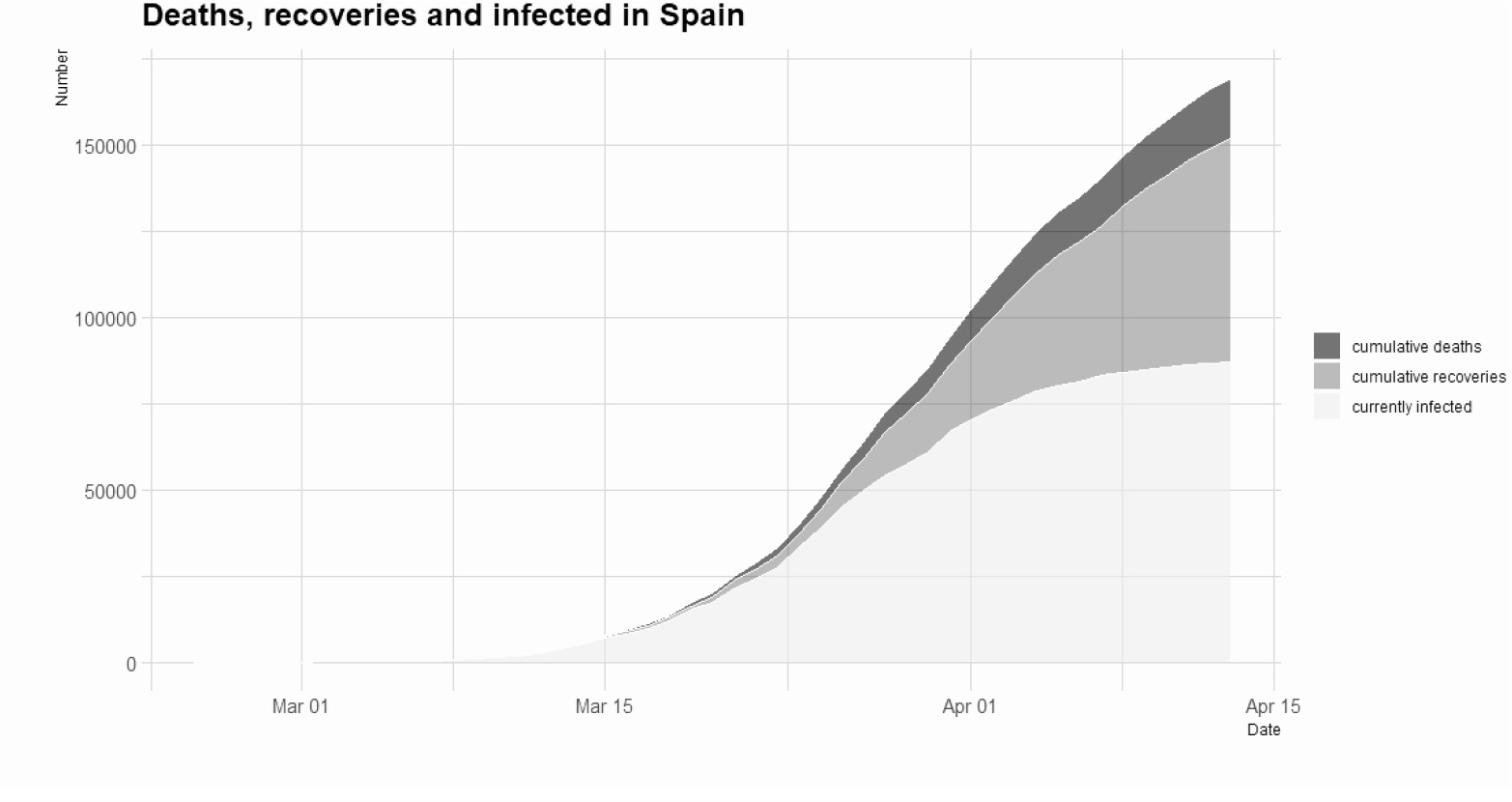
Number of cumulative infected, recovered and dead in Spain

### 5.3 Descriptive analysis

Figure 16 shows the descriptive statistics for Spanish daily infection speed for the period 24 February - 16 May 2020. The mean infection speed (Figure 16a) experiences a smooth and rapid increase during the first month of the observation period, peaking on 26 March, around two weeks after the beginning of nationwide lockdown. After reaching its peak, the mean infection speed begins to decrease volatilely to a level of around 0.002 on 30 April, 35 days after the peak. After this point there is no obvious decreasing trend, however the mean infection speed continues to fluctuate for the remainder of the period. The median (Figure 16a) follows a similar pattern to the mean however is generally lower, again indicating a slight right skewness in daily infection speed distributions. The standard deviation of infection speed (Figure 16b) behaves similarly to the mean and median, however is more volatile than both statistics throughout the period. Skewness (Figure 16c) and kurtosis (Figure 16d) follow similar patterns with their peaks coinciding. Both values are almost always larger than 1 before March 26, the date with greatest mean and standard deviation values. After this point, skewness and kurtosis begin to fluctuate about 1, with a sudden peak on 10 May due to a 10 fold increase in the number of new infected cases in Catalonia that day. Skewness fluctuates between 0 and 4, however for the majority of the period fluctuation between 0 and 2.5 is observed, whilst kurtosis values are almost always positive. Selection of a heavy tailed distribution for infection speed is therefore appropriate in the Spanish case. The apparent change in all four plots on 26 March coincides with the banning of all non-essential activities two days later. The obvious decrease in the mean infection speed after this point supports suggestion of the positive effect of this outbreak control measure on the infection speed in Spain.

**Figure 16.**
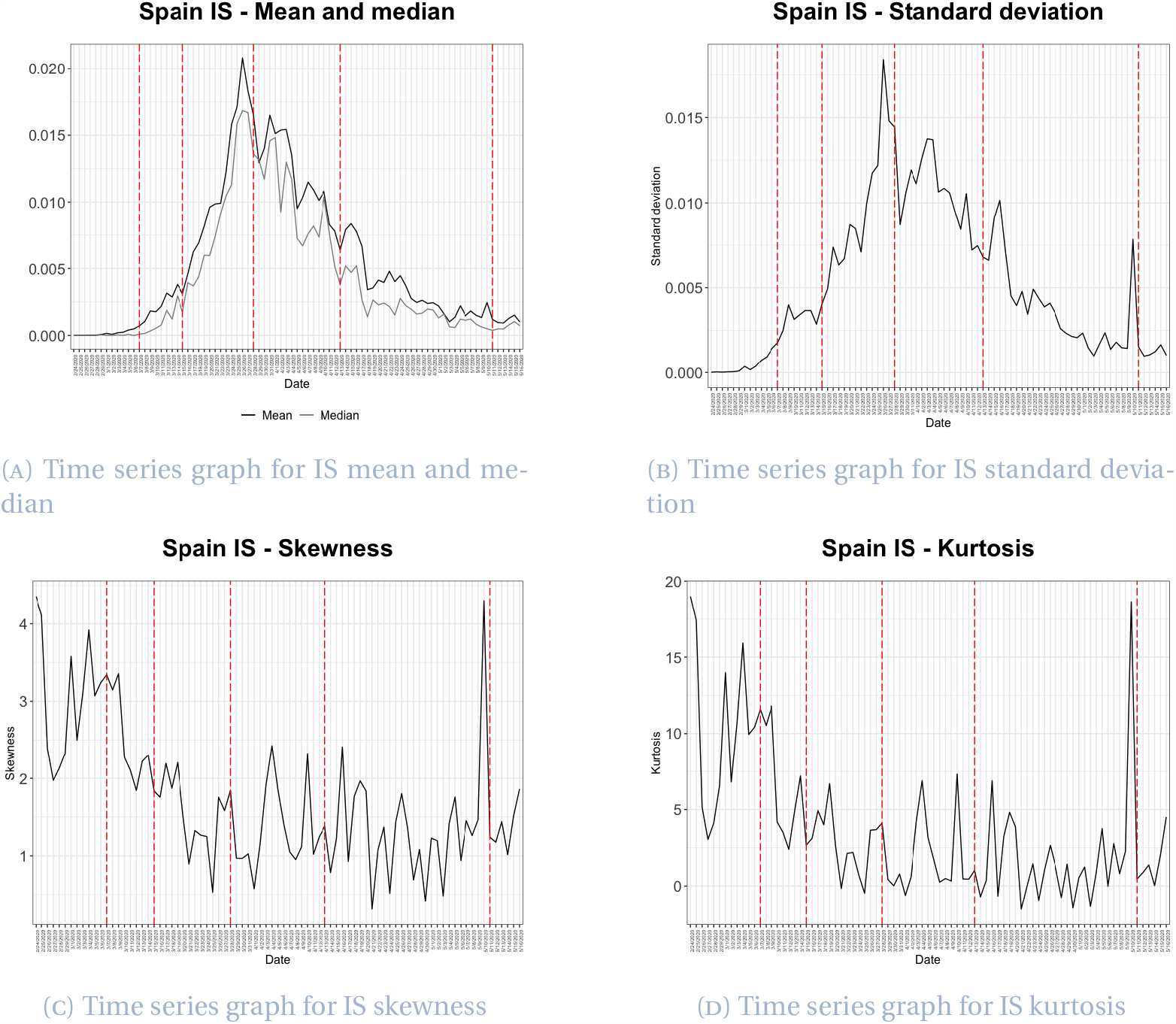
Descriptive statistics for Spanish infection speed

Descriptive statistics for the Spanish fatality ratio are shown in Figure 17 for the interval 8 March - 16 May 2020. Fluctuating for a short period until 20 March, almost one week after the declaration of the nationwide state of emergency and the beginning of lockdown, the mean and median fatality ratio (Figure 17a) begin to increase steadily after this point, stabilising at the beginning of May, approximately six weeks later. The median surpasses the mean on 18 April and remains slightly higher for the remainder of the period. This indicates that the fatality ratio distribution is right skewed until mid April and slightly left skewed afterwards. This distribution change is also observed in Figure 17c, where skewness becomes negative on 10 April. The clear change in distribution approximately one month after the announcement of nationwide lockdown, may highlight the period of delay between imposition and effect. Skewness and kurtosis (Figure 17d) experience perturbations in March despite following a clear downwards trend. Both skewness and kurtosis become stable around the middle of April, with values close to zero. This suggests a symmetric distribution is appropriate for this period. The standard deviation of the fatality ratio (Figure 17b) experiences a significant drop at the very beginning of the observation period, decreasing from 0.10 to 0.032, before beginning a gradual and smooth increase on 14 March, approaching 0.041 towards the end of the period, with all fluctuations smoothed.

**Figure 17.**
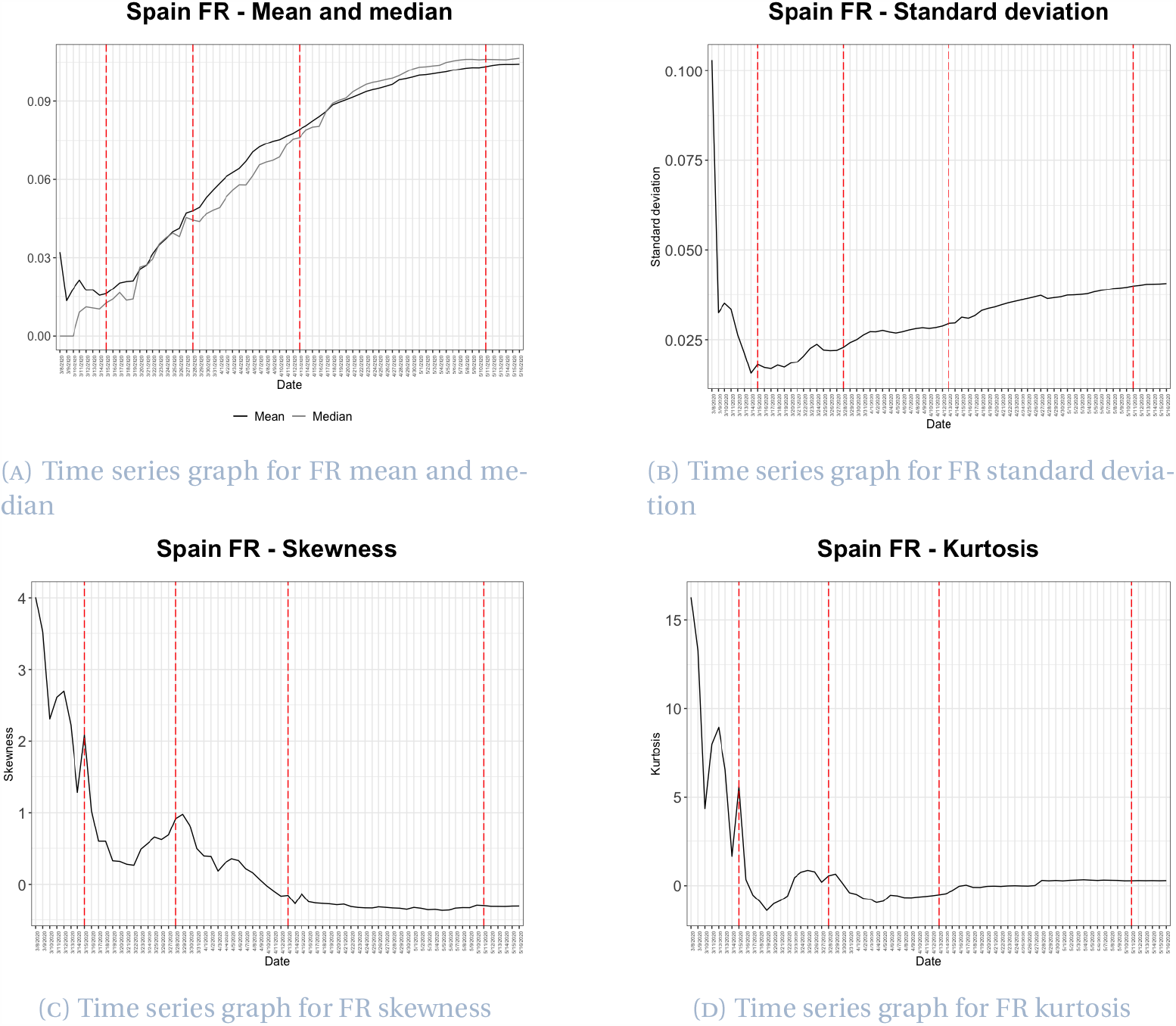
Descriptive statistics for Spanish fatality rate

### 5.4 Distribution fitting

#### Infection speed

The generalised Pareto distribution is selected as the best fitting distribution for Spanish infection speed over the period 26 February - 16 May. This distribution outperforms over all 60 alternative distributions on 52 days of the 82 day period. The probability density function of the generalised Pareto distribution is given in Equation (4.1).

Parameter values for the generalised Pareto distribution for Spanish infection speed are plotted in Figure 18. The parameter *k* is largely positive between 26 February (the date of the first community transmission) and 19 March (four days after nationwide lockdown), before fluctuating with some severity around zero for the remainder of the period. A final peak in the value of *k* is observed on 10 May, one day after the ending of the state of emergency and as discussed in relation to Figure 16, the day of the increase in Catalonian cases. This 10 May peak is however only visible in *k*. The parameter falls back to a value close to zero after this date. For the first 10 days of observation *µ* is stable, the parameter then behaves extremely volatilely throughout the first month of lockdown. Continuing with fluctuations after this point, *µ* gradually becomes increasingly stable and approaches zero from below. The third parameter, *σ*, increases rapidly during the first month of the observation period, and after reaching its peak on 25 March, three days before the banning of all non-essential activities, becomes very volatile and follows a downward trend. With a change in all three parameters observed on 25 March, we observe indications of the effectiveness of lockdown measures after ten days. This observation is paralleled in the infection speed time series’ in Figures 16a and 16b, which peak on 26 March.

**Figure 18.**
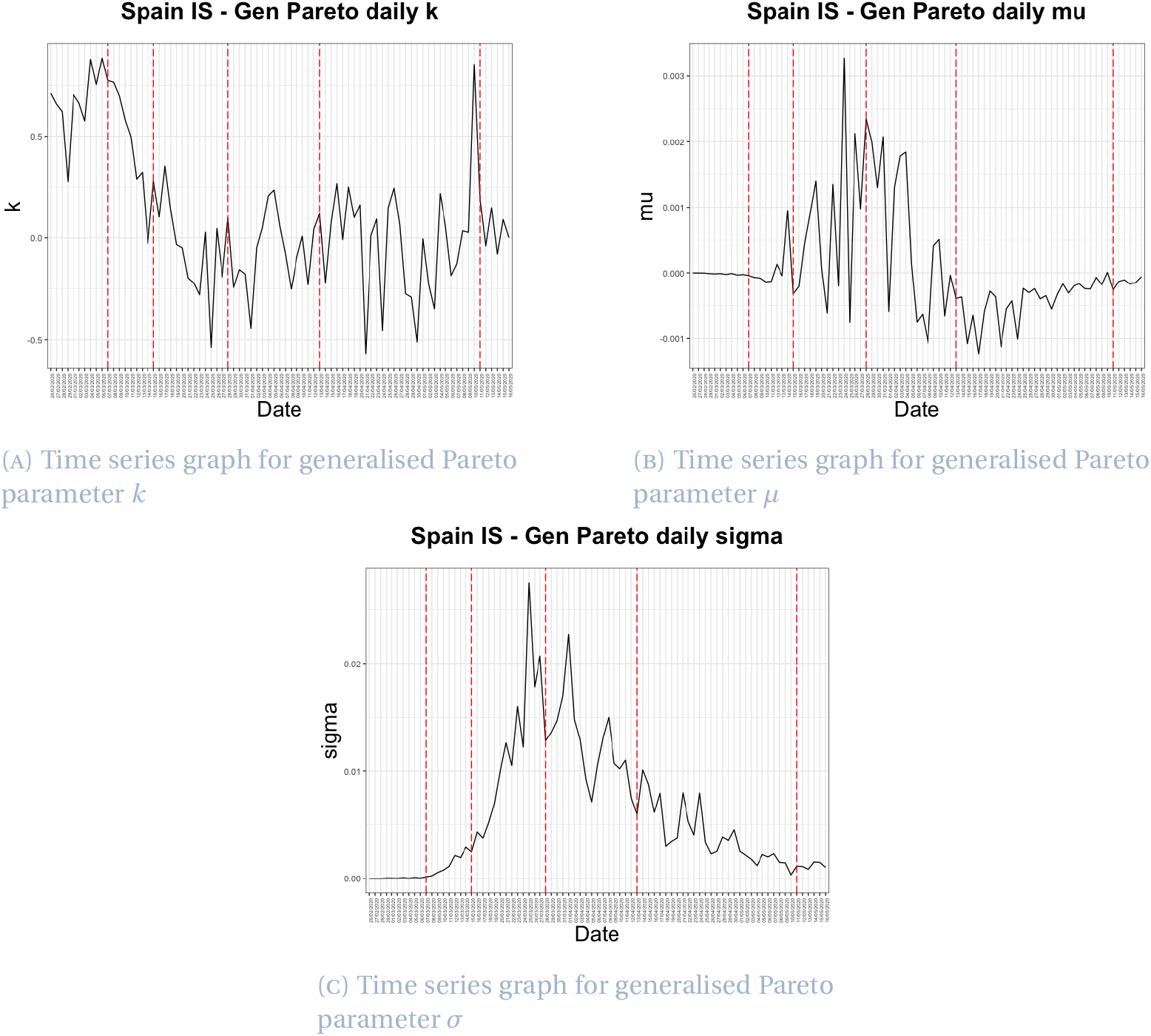
Time series for Spain infection speed generalised Pareto parameters

#### Fatality ratio

After fitting distributions throughout the period 11 March - 16 May, we find the Cauchy distribution to outperform on 30 of 67 days, and so select this distribution as the best fitting distribution. The probability density function of the Cauchy distribution is given by

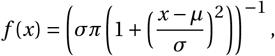

where *µ* is the location parameter and *σ* is the scale parameter (*σ >* 0).

Daily values of the two parameters of the Cauchy distribution for fatality ratio are plotted in Figure 19. The parameter *µ* increases steadily throughout the period. The time series for *µ* is smooth particularly after 29 March, however only minor fluctuations occur prior to this date. Values rise from 0.0059 to 0.10 across the period and reflect the mean and median of the Spanish fatality ratio data. Since *µ* becomes stable after crossing 0.1, it is possible to assume the fatality ratio would not change significantly in the future, in the event there was no second wave of the outbreak. The slowing of the rate of change of the fatality ratio parameter *µ* appears to begin one month into the observation period, on 11 April. This date is approximately one month after the declaration of nationwide lockdown and two weeks after the banning of all non-essential activities. The second parameter, *σ*, again follows an increasing trend however is unstable until 20 April, after which point the volatility in the daily parameter values decreases. The timing of the changes in both parameter values suggests a stabilising of the fatality ratio as a result of lockdown measures which takes approximately one month to come into effect. The largest changes in the *σ* time series appear to coincide with the highlighted dates, which are those of greatest significance in regard to the implementation of government measures, inferring the sensitivity of the distribution to certain changes in the outbreak environment. Changes in the *µ* and *σ* plots on 11 and 20 April, respectively, fall in line with the change in distribution implied by the mean, median and skewness plots in Figure 17. However, the parameter change is much less significant.

**Figure 19.**
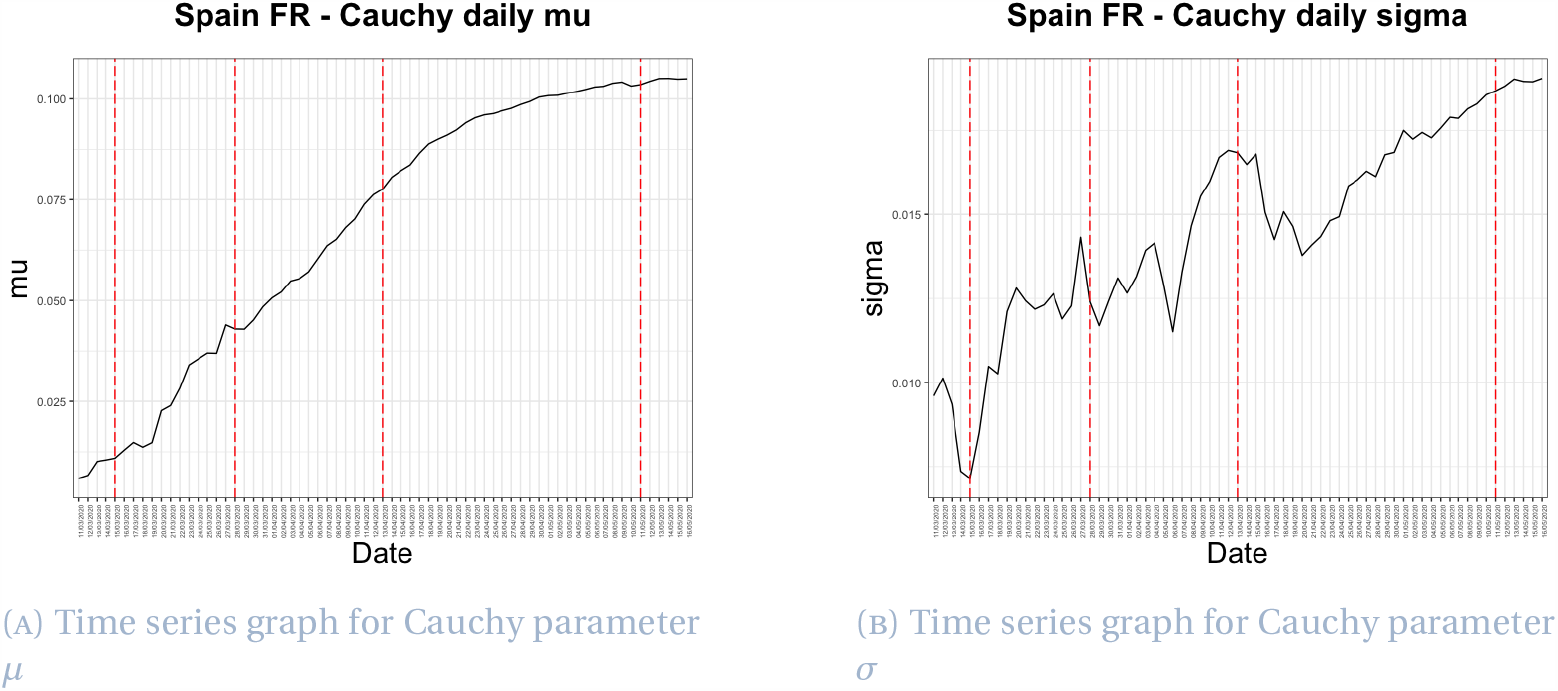
Time series for Spain fatality ratio Cauchy parameters

## 6 Concluding remarks

The previous three sections provide detailed overviews of the changing severity of the COVID-19 outbreak in Hubei, Italy and Spain, presenting evidence for the significance of certain government measures and particularly lockdown restrictions through observation of their death rate, infection speed and fatality ratio. Infection status is generally more certain in comparison to determining an individual’s cause of death, particularly with the varying methods for counting COVID-19 fatalities. The accuracy of all three severity measures however, relies upon the sufficient testing of the populations under consideration.

As of 17 September 2020, 215 countries, areas and territories had reported cases of COVID-19. In particular, the WHO reported 29,737,453 cases worldwide, with 937,391 deaths. The steady decline in the number of infected cases as of mid-February observed in China, the first reporting site of the novel coronavirus, may be attributable to extreme measures taken by the government in order to contain the virus, with the majority of new cases imported from abroad by mid July and only occasional cases of domestic transmission. At this time, new daily cases were reported to be approximately 200 and 700 in Italy and Spain, respectively, a significant reduction in comparison to the equivalent values in March and April.

The analysis presented in this paper supports suggestion that the imposition of lockdown has a positive effect on all three severity measures considered. In general, the mean, median and standard deviation of the infection speed and death rate in each of the three territories experience an increasing and decreasing trend (with varying severity) which aligns with the period of lockdown. Death rate decreases and stabilisation reflect improvements in medical treatments in addition to better management of COVID-19 infections. Whilst a decreasing average is intuitive when considering lockdown to be a positive measure, a decline in the standard deviation reflects the evening out of the prevalence and severity of the infection across regions within each territory. This increased spread alleviates pressure on medical demand and government support during the outbreak. Considering for example the case in Italy, a high number of infections in the Northern region at the beginning of the outbreak induced a high standard deviation of the death rate, with the majority of Italy reporting no infections at this point in time. As the outbreak spread throughout the country, the variance in both infection speed and death rate stabilised. Occurring simultaneously to the stabilisation of the mean and median, this suggests the outbreak was becoming increasingly controlled.

Discussion of descriptive statistics and the distribution fitting analysis provides an understanding of the pandemic environment during the first quarter of its outbreak. Although we cannot draw firm conclusions in regard to the timeliness of lockdown measures, and in particular, the impact of a shift in the period of lockdown on the severity of the outbreak, due to the available data and the chosen approach, the positive effect of the introduction of lockdown on stabilising infection speed, death rate and fatality ratio is clear in all three cases. This paper provides thorough documentation of the initial stages of the pandemic in Hubei, Italy and Spain, creating a reference for the manner in which disease spread changes in relation to government measures, in the event of the occurrence of any outbreak of a similar nature in the future.

## Data Availability

The R package, nCov2019, developed by Yu (202), provides direct access to real-time epidemiological data on the outbreak.
Yu, G. (2020). nCov2019: An R package with real-time data, historical data and Shiny app.

## Acknowledgments

M.C. Boado-Penas is grateful for the financial assistance received from the Spanish Ministry of the Economy and Competitiveness [project ECO2015-65826-P]. The research of J. Eisenberg was funded by the Austrian Science Fund (FWF), Project number V 603-N35.

## Footnotes

1 After the initial reporting of the outbreak in Wuhan, Deng and Peng [3], Yang et al. [15] and Sohrabi et al. [10], China quickly entered a state of fighting against the new coronavirus based on their experience in the use of suppression policies during the SARS-CoV epidemic. Individual behaviour has been crucial in controlling the spread of COVID-19. As a result of the suppression measures taken by the government, on January 23rd 2020 Wuhan was quarantined and movement was restricted across Hubei province, affecting 50 million people. All public transportation was sealed off within China while outside of China aviation restrictions were applied and several countries initiated temperature and symptom screening protocols towards Chinese citizens. In Wuhan, two new hospitals were built in two weeks in early February to treat coronavirus patients.

2 “Coronavirus: China’s first confirmed Covid-19 case traced back to November 17”, Ma, J., *South China Morning Post*, March 2020, https://www.scmp.com/news/china/society/article/3074991/coronavirus-chinas-first-confirmed-covid-19-case-traced-back?onboard=true

3 “Tracing the New Coronavirus gene sequencing: when did the alarm sound”, Gao, Y. and Peng, Y. and Yang, R. and Feng, Y. and Ma, D., *Caixin Global*, February 2020, https://web.archive.org/web/20200227094018/http://china.caixin.com/2020-02-26/101520972.html

4 “Li Wenliang: Coronavirus kills Chinese whistleblower doctor”, McDonell, S., *BBC News*, February 2020, https://www.bbc.com/news/world-asia-china-51403795

5 “Wuhan unexplained pneumonia has been isolated test results will be announced as soon as available”, Yang, X., *Yicai Global*, December 2019, https://m.yicai.com/news/100451932.html

6 “Wuhan Municipal Health Commission’s message about our city’s present pneumonia situation”, Wuhan Municipal Health Commission, December 2019, https://web.archive.org/web/2020010921541 3/http://wjw.wuhan.gov.cn/front/web/showDetail/2019123108989

7 “Pneumonia of unknown cause – China”, World Health Organization, January 2020, https://www.who.int/csr/don/05-january-2020-pneumonia-of-unkown-cause-china/en/

8 “8 Legally Sanctioned for Spreading False Information on the ‘Wuhan Viral Pneumonia”‘, Liao, J. and Feng, G., *Xinhua Net*, January 2020, http://www.xinhuanet.com/2020-01/01/c_1125412773.htm

9 “China releases genetic data on new coronavirus, now deadly”, Schnirring, L., *CIDRAP News*, January 2020, https://www.cidrap.umn.edu/news-perspective/2020/01/china-releases-genetic-data-new-coronavirus-now-deadly

10 “China Reports First Death From New Virus”, Qin, A. and Hernández, J. C., *The New York Times*, January 2020, https://www.nytimes.com/2020/01/10/world/asia/china-virus-wuhan-death.html

11 “China confirms human-to-human transmission of coronavirus”, Kuo, L, *The Guardian*, January 2020, https://www.theguardian.com/world/2020/jan/20/coronavirus-spreads-to-beijing-as-china-confirms-new-cases

12 “Coronavirus hospital set to open in Wuhan with 1,400 military medical staff”, Wu, W., *South China Morning Post*, February 2020, https://www.scmp.com/news/china/politics/article/3048592/coronavirus-hospital-set-open-wuhan-1400-military-medical-staff

13 “How China Built Two Coronavirus Hospitals in Just Over a Week”, Wang, J. and Zhu, E. and Umlauf, T., *The Wall Street Journal*, February 2020, https://www.wsj.com/articles/how-china-can-build-a-coronavirus-hospital-in-10-days-11580397751

14 “Coronavirus: China reports 254 deaths in a day as cases surge after including clinically diagnosed patients”, Zhou, C. and Choy, G. and Zhuang, P., *South China Morning Post*, February 2020, https://www.scmp.com/news/china/society/article/3050354/coronavirus-hubei-province-reports-sharp-spike-new-confirmed

15 “China to lift travel restrictions in Hubei after months of coronavirus lockdown”, Davidson, H., *The Guardian*, March 2020, https://www.theguardian.com/world/2020/mar/24/china-to-lift-travel-restrictions-in-hubei-after-months-of-coronavirus-lockdown

16 “China will lift lockdown on Wuhan, the epicenter of coronavirus outbreak, on April 8”, O’Kane, C., *CBS News*, March 2020, https://www.cbsnews.com/news/coronavirus-update-china-lift-lockdown-wuhan-april-8-epicenter-quarantine/

17 “China’s Wuhan raises Covid-19 death toll by 50%, citing early lapses”, Chen, Y. and Goh, B., *Reuters*, April 2020, https://www.thejakartapost.com/news/2020/04/17/chinas-wuhan-raises-coronavirus-death-toll-by-50-citing-early-lapses.html

18 “Coronavirus: China outbreak city Wuhan raises death toll by 50%”, McDonell, S., *BBC News*, April 2020, https://www.bbc.com/news/world-asia-china-52321529

19 “Spain daily coronavirus death toll lowest in weeks: Live updates”, Rasheed, Z. and Stepansky, J. and Alla-houm, R., *Al Jazeera*, April 2020, https://web.archive.org/web/20200427041920https://www.aljazeera.com/news/2020/04/global-coronavirus-deaths-pass-200000-live-updates-200425232324631.html

20 CNHC holds the historical statistics for the 34 provinces and special districts in China.

21 Dingxiangyuan continuously aggregates data from provincial and city health agencies and the CNHC [4].

22 This GitHub repository [14] derives data from the literature Huang et al. [5] for December 1, 2019, to January 10, 2020, after which it relies on the Chinese news aggregator Toutiao API.

23 “Coronavirus, primi due casi in Italia”, Severgnini, C., *Corriere della Sera*, January 2020, https://www.corriere.it/cronache/20_gennaio_30/coronavirus-italia-corona-9d6dc436-4343-11ea-bdc8-faf1f56f19b7.shtml

24 “Italy declares state of emergency”, Borghese, L., *CNN*, January 2020, https://edition.cnn.com/asia/live-news/coronavirus-outbreak-01-31-20-intl-hnk/h_ed756d2007470c7fb4eeb4492bafabf5

25 “Factbox: Latest on coronavirus spreading in China and beyond”, Daniel, A. C. and Nissi, M. and Kalluvila, S., *Reuters*, February 2020, https://uk.reuters.com/article/us-china-health-latest-factbox/factbox-latest-on-coronavirus-spreading-in-china-and-beyond-idUKKBN20E1BF

26 “Italian towns on lockdown after 2 virus deaths, clusters”, Bruno, L. and Winfield, N., *CTV News*, February 2020, https://www.ctvnews.ca/world/italian-towns-on-lockdown-after-2-virus-deaths-clusters-1.4823230

27 “What towns in Italy are on lockdown because of coronavirus?”, Paul, A., *Metro*, February 2020, https://metro.co.uk/2020/02/25/towns-italy-lockdown-coronavirus-12298246/

28 “Italy under lockdown: ‘My town is shocked and scared”‘, Johnson, M. and Ghiglione, D, *Financial Times*, February 2020, https://www.ft.com/content/8f34c332-5947-11ea-abe5-8e03987b7b20

29 “Coronavirus, scuole chiuse in tutta Italia fino a metà marzo”, Guerzoni, M. and Santarpia, V., *Corriere della Sera*, March 2020, https://www.corriere.it/scuola/20_marzo_04/coronavirus-scuole-chiuse-tutta-italia-decisione-governo-entro-stasera-e7ba0614.-5e12-11ea-8e26-25d9a5210d01.shtml

30 “Italy orders closure of all schools and universities due to coronavirus”, Giuffrida, A. and Tondo, L. and Beaumont, P., *The Guardian*, March 2020, https://www.theguardian.com/world/2020/mar/04/italy-orders-closure-of-schools-and-universities-due-to-coronavirus

31 “Italy quarantines 16 million people over coronavirus fears”, Bertelli, M., *Al Jazeera*, March 2020, https://www.aljazeera.com/news/2020/03/italy-quarantines-quarter-population-fight-coronavirus-200308071832617.html

32 “Italy extends emergency measures nationwide”, Archeta, K., *COVID-19 World News*, March 2020, https://covid19data.com/2020/03/10/italy-extends-emergency-measures-nationwide/

33 “Coronavirus: Italy bans any movement inside country as toll nears 5,500”, Giuffrida, A. and Safi, M. and Farrer, M., *The Guardian*, March 2020, https://www.theguardian.com/world/2020/mar/22/italian-pm-warns-of-worst-crisis-since-ww2-as-coronavirus-deaths-leap-by-almost-800

34 “Italy records lowest coronavirus death toll for a week”, Henley, J., *The Guardian*, April 2020, https://www.theguardian.com/world/2020/apr/01/italy-extends-lockdown-amid-signs-coronavirus-infection-rate-is-easing

35 “L’Italia chiude i porti”, Salvatori, P., *HuffPost*, April 2020, https://www.huffingtonpost.it/entry/litalia-chiude-i-porti_it_5e8d89f2c5b6e1d10a6c2671?yhj&utm_hp_ref=it-homepage

36 “Italy Goverment Set to Extend Virus Lockdown to May 3: Ansa”, Follain, J., *Bloomberg*, April 2020, https://www.bloomberg.com/news/articles/2020-04-10/italy-goverment-set-to-extend-virus-lockdown-to-may-3-ansa

37 “Italy PM extends virus lockdown, says euro zone rescue plan inadequate”, Jones, G. and Amante, A., *Reuters*, April 2020, https://www.reuters.com/article/us-health-coronavirus-italy-conte/italy-extends-coronavirus-lockdown-until-may-3-prime-minister-idUSKCN21S1YL

38 “Italy sees first fall of active coronavirus cases”, Mayberry, K. and Stepansky, J. and Varshalomidze, T., *Al Jazeera*, April 2020, https://www.aljazeera.com/news/2020/04/coronavirus-deaths-exceed-40000-live-updates-200419233722851.html

39 “Discorso di Conte in conferenza stampa, le riaperture dal 18 maggio: “Corriamo un rischio calcolato”“, Severgnini, C., *Corriere della Sera*, May 2020, https://www.corriere.it/politica/20_maggio_16/discorso-conte-conferenza-stampa-oggi-decreto-18-maggio-1e810142-9785-11ea-ba09-20ae073bed63.shtml

40 “Spain confirms first case of Wuhan coronavirus”, Linde, P., *El País*, February 2020, https://english.elpais.com/international/2020-02-03/spain-confirms-first-case-of-wuhan-coronavirus.html?rel=listapoyo

41 “First confirmed coronavirus case in Spain in La Gomera, Canary Islands”, Bilbatua, J., *Outbreak News Today*, February 2020, http://outbreaknewstoday.com/first-confirmed-coronavirus-case-in-spain-in-la-gomera-canary-islands-20628/

42 “Spanish town faces police lockdown to contain coronavirus”, Jones, S., *The Guardian*, March 2020, https://www.theguardian.com/world/2020/mar/07/spanish-town-faces-police-lockdown-to-contain-coronavirus

43 “España prohíbe todos los vuelos directos procedentes de Italia hasta el 25 de marzo”, Vega, M., *El Español*, March 2020, https://www.elespanol.com/invertia/empresas/20200310/espana-prohibe-vuelos-directos-italia-marzo/473703280_0.html

44 “Spanish government declares state of alarm”, Cué, C. E. and Pérez, C., *El País*, March 2020, https://english.elpais.com/politics/2020-03-13/spanish-government-declares-state-of-alarm-in-bid-to-combat-coronavirus-spread.html?rel=listapoyo

45 “El Gobierno informa de que es la única autoridad en toda España, limita los desplazamientos y cierra comercios”, Cué, C. E., *El País*, March 2020, https://elpais.com/espana/2020-03-14/el-gobierno-prohibe-todos-los-viajes-que-no-sean-de-fuerza-mayor.html

46 “Spain to impose nationwide lockdown - El Mundo”, Melander, I. and Jones, J., *Reuters*, March 2020, https://nationalpost.com/pmn/health-pmn/spain-to-impose-nationwide-lockdown-el-mundo

47 “Spain may be a week ahead of the U.S. in its coronavirus quarantine: Here’s what you can learn from its experience”, Kollmeyer, B., *MarketWatch*, March 2020, https://www.marketwatch.com/story/spain-may-be-a-week-ahead-of-the-us-in-its-coronavirus-quarantine-heres-what-you-can-learn-from-its-experience-2020-03-13

48 “Spain orders nationwide lockdown to battle coronavirus”, Jones, S., *The Guardian*, March 2020, https://www.theguardian.com/world/2020/mar/14/spain-government-set-to-order-nationwide-coronavirus-lockdown

49 “Spanish government tightens lockdown to include all non-essential workers”, Marcos, J., *El País*, March 2020, https://english.elpais.com/politics/2020-03-28/spanish-government-tightens-lockdown-to-include-all-non-essential-workers.html?rel=listapoyo

50 “Spain orders non-essential workers stay home for two weeks”, Jones, S., *The Guardian*, March 2020, https://www.theguardian.com/world/2020/mar/28/covid-19-may-be-peaking-in-parts-of-spain-says-official

51 “Congress backs PM’s request to extend state of alarm in Spain until April 26, with a further 15 days likely”, Díez, A. and Casqueiro, J., *El País*, April 2020, https://english.elpais.com/politics/2020-04-10/congress-backs-pms-request-to-extend-confinement-measures-in-spain-until-april-26-with-a-further-15-days-likely.html?rel=listapoyo

52 “Spain extends state of emergency until May 9, prolonging lockdown to 8 weeks”, Alberti, M. and Rebaza, C. and Formanek, I. and Goodman, A. and Tejera, I., *CNN*, April 2020, https://edition.cnn.com/world/live-news/coronavirus-pandemic-04-23-20-intl/h_5757522b8521fdeda9a5c6e914a7b7c6

53 “Spain’s Government approves territories that can move to Phase 1 of de-escalation lifting restrictions for 51% of its population – here’s the complete list”, Sappal, P., *Euro Weekly News*, May 2020, https://www.euroweeklynews.com/2020/05/08/breaking-news-spains-government-approves-territories-that-can-move-to-phase-1-of-deescalation-lifting-restrictions-for-51-of-its-population-heres-the-complete-list/

54 “El mapa de la desescalada: las provincias y territorios que pasan de fase el lunes 25 de mayo”, Hernández, Q., *La Sexta*, May 2020, https://www.lasexta.com/noticias/nacional/mapa-desescalada-provincias-territorios-que-pasan-fase-lunes-mayo_202005225ec7cb0aea9d4700011bbed9.html

55 “Toda España estará el lunes en Fase 3 menos Madrid, algunas provincias de Castilla y León y zonas de Barcelona y Lleida”, León, A., *RTVE*, June 2020, https://www.rtve.es/noticias/20200612/toda-espana-estara-lunes-fase-3-menos-madrid-algunas-provincias-castilla-leon-barcelona-lleida/2017322.shtml

56 “As Spain enters the ‘new normality’ on Sunday, what will change?”, Hunter, S., *El País*, June 2020, https://english.elpais.com/society/2020-06-20/as-all-of-spain-enters-the-new-normality-on-sunday-what-will-change.html

57 “Así es la nueva normalidad”, Villahizán, J., *El Día de Valladolid*, June 2020, https://www.eldiadevalladolid.com/Noticia/Z5EA0E550-C084-4FC6-AAB1B772D7539471/202006/As%C3%AD-es-la-nueva-normalidad

58 “Actualización no. 77. Enfermedad por el coronavirus (COVID-19). 16.04.2020”, Coordination Center for Health Alerts and Emergencies, April 2020, https://www.mscbs.gob.es/profesionales/saludPublica/ccayes/alertasActual/nCov-China/documentos/Actualizacion_77_COVID-19.pdf

